# MSGene: Derivation and validation of a multistate model for lifetime risk of coronary artery disease using genetic risk and the electronic health record

**DOI:** 10.1101/2023.11.08.23298229

**Authors:** Sarah M. Urbut, Ming Wai Yeung, Shaan Khurshid, So Mi Jemma Cho, Art Schuermans, Jakob German, Kodi Taraszka, Akl C. Fahed, Patrick Ellinor, Ludovic Trinquart, Giovanni Parmigiani, Alexander Gusev, Pradeep Natarajan

## Abstract

Currently, coronary artery disease (CAD) is the leading cause of death among adults worldwide. Accurate risk stratification can support optimal lifetime prevention. We designed a novel and general multistate model (MSGene) to estimate age-specific transitions across 10 cardiometabolic states, dependent on clinical covariates and a CAD polygenic risk score. MSGene supports decision making about CAD prevention related to any of these states. We analyzed longitudinal data from 480,638 UK Biobank participants and compared predicted lifetime risk with the 30-year Framingham risk score. MSGene improved discrimination (C-index 0.71 vs 0.66), age of high-risk detection (C-index 0.73 vs 0.52), and overall prediction (RMSE 1.1% vs 10.9%), with external validation. We also used MSGene to refine estimates of lifetime absolute risk reduction from statin initiation. Our findings underscore the potential public health value of our novel multistate model for accurate lifetime CAD risk estimation using clinical factors and increasingly available genetics.

## Introduction

Coronary artery disease (CAD), remains the leading cause of morbidity and mortality worldwide.^1^ Estimating an individual’s risk of developing CAD over the lifetime is essential for timely and effective prevention and intervention.^2–5^ Traditional risk prediction models, such as the Pooled Cohort Equations (PCE) 10-year risk score, have guided clinical decisions and preventive strategies; however, these models come with inherent limitations.^6–8^ A 30-year or 10-year window provides only a fixed, albeit extended, snapshot of risk. It neither captures the entirety of an individual’s lifetime risk nor provides dynamic, age-specific insights beyond these arbitrary periods. Most importantly, there is a growing need for models capable of both recognizing undertreated younger patients while reducing over-estimation in older patients.^7,9,10^

Current guidelines^9,11,12^ recommend the consideration of primordial risk factors in risk-stratifying patients, and call for better methods of estimating lifetime risk. Recent evidence suggests that lifetime risk assessment provides a more comprehensive picture of an individual’s propensity for developing CAD across time.^13,14^ Traditional factors in combination with genomic risk can confer a disproportionately elevated risk for CAD in the long term.^2,15–17^ Focusing on lifetime risk allows for more effective patient counseling, tailored preventive measures, and earlier interventions that may delay or prevent the onset of CAD altogether.^18,19^

Because of the multifactorial nature of CAD, there is an increasing need for continuously updated, dynamic and individualized CAD risk predictions that span a patient’s entire life.^2,14,20^ Such risk prediction models could improve the identification of undertreated younger patients while avoiding risk over-estimation in older patients.^7,9,10^ Understanding risk from this perspective allows for more informed and timely interventions, potentially even before the conventional risk windows are applicable.

Here, we introduce the MSGene model — a multistate model designed to predict the lifetime risk of CAD, conditional on both time-fixed and time-dependent variables. Multistate models allow for the estimation of the risk of an individual transitioning between health states^21–25^ through flexible estimation of conditional probabilities by modeling the transitions between states over time. By modeling the different health states simultaneously, they naturally account for competing risks.

MSGene is capable of modeling the dynamic transitions from risk factor states to CAD with age-specific coefficients. Critically, our approach differs from a traditional Markov-based multistate model^21,22^ by extending our model to the time inhomogeneous case and allowing our transitions to vary with age, and also from traditional Cox models by allowing for non-proportional hazards.

In the current study, we develop and validate the MSGene model. We evaluate the performance compared to the traditionally employed Framingham 30-year^26^ and PCE 10-year^5,6^ models. We then estimate the potential ability of MSGene to reduce CAD events by guiding timely initiation of statin therapy and demonstrate the benefit of a multistate framework to incorporate dynamic changes in treatment decisions for unique patient profiles.

## Results

### Novel multistate model with time-dependent transitions

We build a novel time-dependent multistate model in which age is the time scale. For each age and current state (**Fig. 1**), we model the one-year probability of transition from state to state as a logistic regression conditional on both time-fixed covariates (sex, CAD-PRS), and time-dependent covariates (smoking, use of anti-hypertensives or statins) (**Methods**). This methodology defines an inhomogenous Markov transition model which can be used to compute the probability of reaching any state of interest during one’s lifetime, among other quantities. Here, to compare our model to existing tools we focus on CAD.

**Figure 1.**
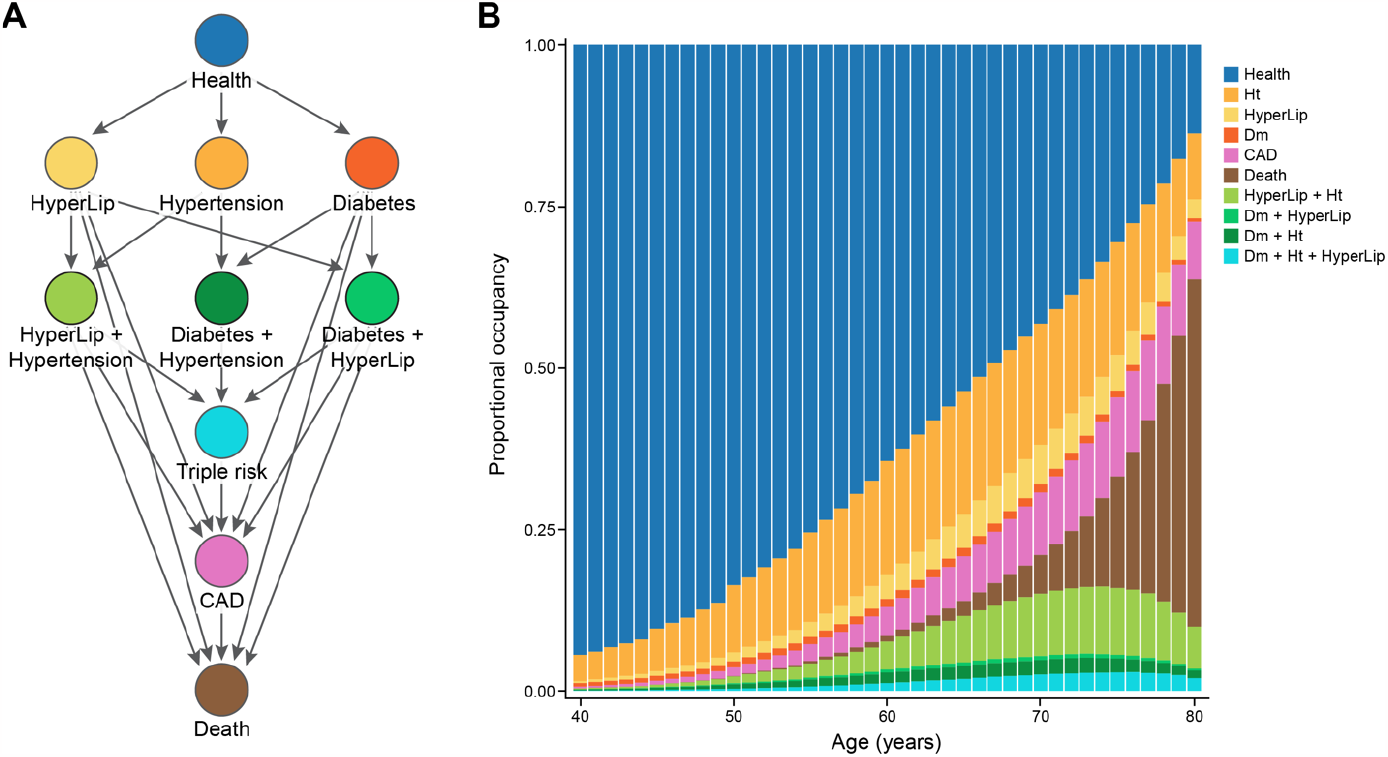
Multistate transitions over time. **A**. We depict the potential one-step transitions in our multistate framework. Per year, an individual can progress from health to single risk factor states, CAD or death. Similarly, an individual can progress from single risk factor states, to double risk factor states, to CAD or death; from double risk factor states, to triple risk factor, CAD or death. **B**. We display the proportional occupancy excluding censored individuals at each state. **CAD:** coronary artery disease, **Ht**: hypertension, **HyperLip:** hyperlipidemia, **Dm:** Type 2 diabetes mellitus.

We use a limited set of covariates (**Methods)** as a result of the variable selection described in **Supp. Table 1**. To improve estimation efficiency, we smooth each set of state to state coefficients across ages using a flexible tricube distance weighted least square local regression^27^ with inverse variance weighting of raw estimate. This allows for the sharing of information across ages in instances in which the number of individuals at a particular transition may be small. We calculate risk under a statin-treated and statin-untreated strategies by imputing trial-imputed relative risk reduction of statin use on each annual age-specific transition (**Methods**). We develop this in the R programming language (4.3.0) and provide detailed code and an interactive application for users.

**Table 1.**
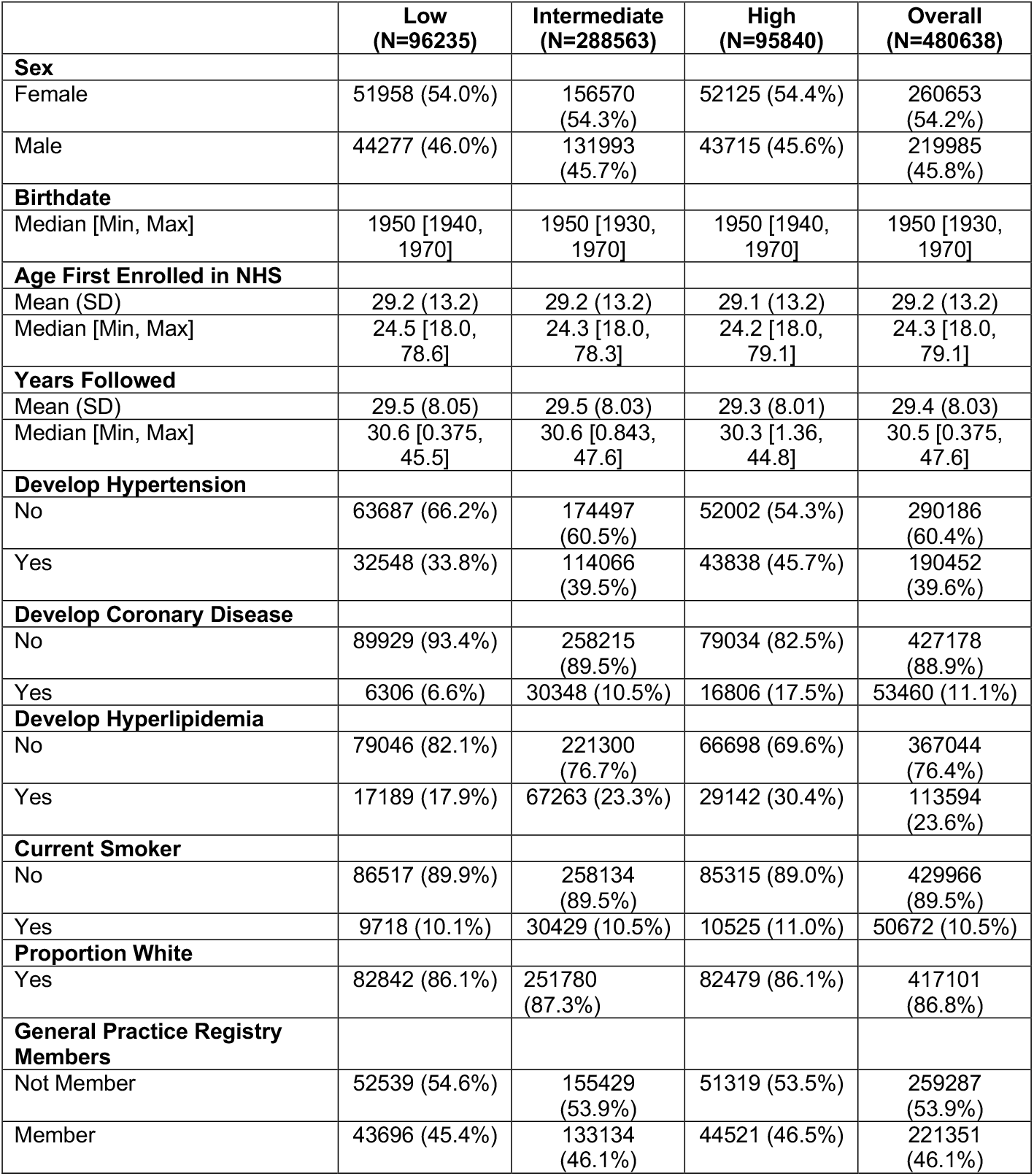
Distribution of overall cohort. We use approximately 80% (385,541) individuals in the training, and 79,119 in the testing set, of which approximately 45% represent members of the general practice primary care data. Of note, low genomic risk connotes individuals in the lowest (<20%) of genomic risk by PRS percentile, intermediate (20–80%) PRS percentile, and high denotes >80% PRS percentile.

### Baseline characteristics

We considered 480,638 individuals: 260653 (54.2%) were female with 43,855 (11.1%) incident coronary artery disease diagnoses (**Table 1**) with a median 29.9 years [22.4–35.1] years of follow-up and median age of first observation in EHR 24.3 [IQR: 18.0, 37.1] after excluding 20,534 who lacked sufficient covariates or had CAD at baseline (**Fig. 2**). MSGene allows for visualization of the proportional representation by risk factor at each age (**Fig. 1**): approximately 39.6% are ultimately diagnosed with hypertension, 23.6% with hyperlipidemia, and 9.9% with Diabetes mellitus (1 or 2). Furthermore, 10.5% report currently smoking and 20.3% began antihypertensive use during the course of our study; 46.1% also contributed to the general practice cohort, and the distribution of risk factors was homogenous between subsets (**Supp. Fig. 1**). We use 80% of our data as training and 20% as testing (**Fig. 2**) for internal cross-validation and to optimize model fit. Accordingly, this divides our data into a training set for model fitting using 384,510 samples and a testing data set of 79,117 unique individuals. We report. the lifetime risk remaining at any age as one minus the product of the complement of the interval age and state-specific transition to CAD probabilities.

**Figure 2.**
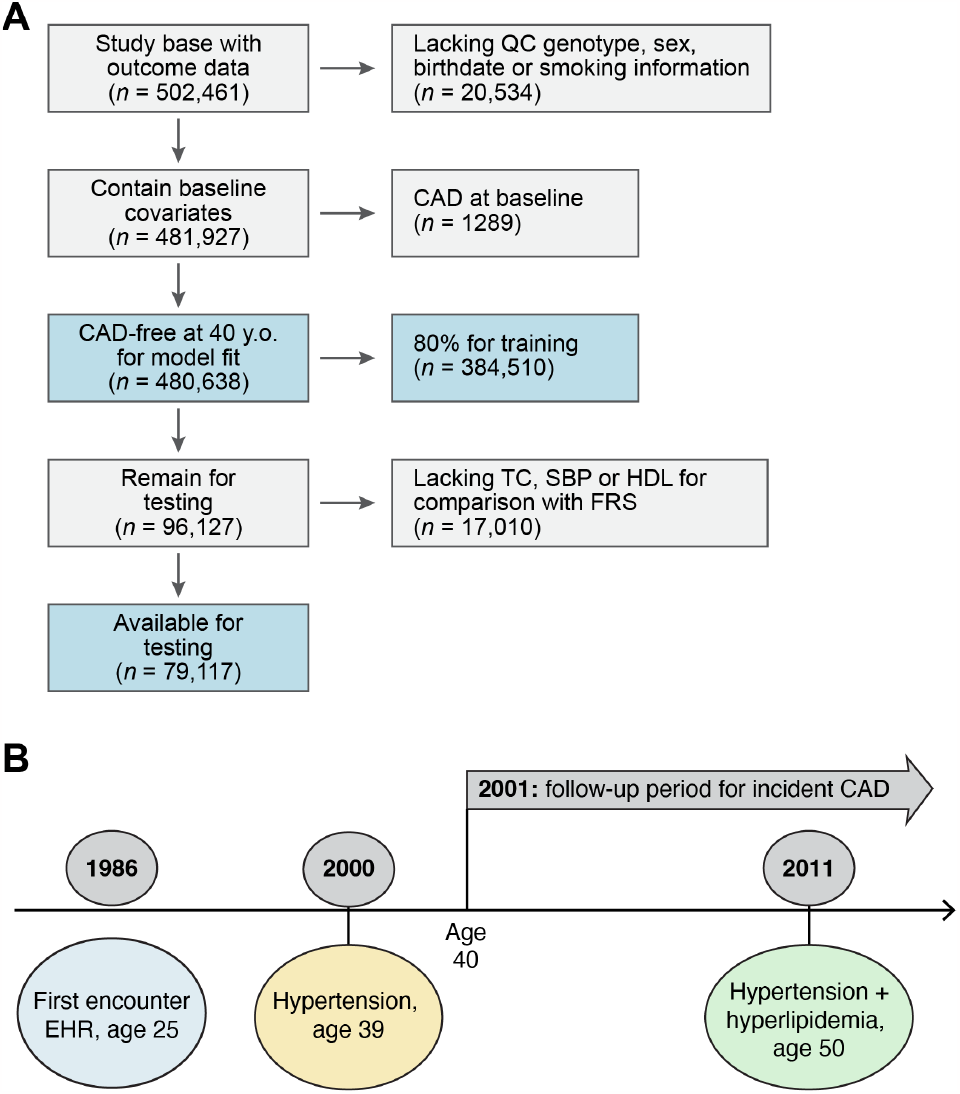
Study overview. **A**. Using the UK Biobank data on half a million participants (54% female) with access to health record from 1940, we harmonize hospitalization, prescription and primary care records from the EHR and train our model on individuals free of CAD at age 40. The UKB required participants to be between ages 40–69 between 2006–2010 for genotyping. In our model, individuals join disease-free in the ‘health’ state and progress to additional states upon censoring. We use 80% of the eligible data for training and the remaining 20% for testing. For the testing subset we require that individuals have variables necessary for computation of FRS30 (and FRS30RC) and the pooled cohort equations, which require laboratory (HDL, TC) and biometric (SBP) measurements. **B**. For a sample patient, we document the construction of our cohort. This individual is first observed in the health record at age 25; he is diagnosed with hypertension at age 39, and begins informing our risk estimation for CAD at age 40 in the hypertensive category. He transitions to the hypertension and hyperlipidemia category at age 50, 25 years after first encounter and 10 years after entering our risk estimation, thus contributing 10 years of data. **TC:** total cholesterol, **SBP**: systolic blood pressure, **HDL**: high-density lipoprotein, **CAD**: coronary artery disease, **FRS30**: Framingham 30 year, **FRS30RC**: Framingham 30 year recalibrated, **PCE:** Pooled cohort equation 10-year risk; **EHR**: electronic health record.

### Modeling transitions

Using our multistate approach, MSGene, we describe the overall state distribution across the lifespan in our cohort, normalizing to exclude censoring at each age (**Fig. 1)**. At age 40 years, 94.4% of individuals are in the healthy category, with 4.1% in the hypertensive category and 0.3% with a diagnosis of CAD. By age 76 years, CAD state occupancy peaks at 12.5% of uncensored individuals, and health is reduced to 27.6% of uncensored individuals. By age 80 years, 7.4% have died.

### Improved detection of early events when compared to 10-year risk

When compared to the PCE, a 10% lifetime threshold using MSGene uniquely identifies 5315 (59.3%) cases versus 123 (1.3%) cases using the 10-year PCE (5% threshold) alone at age 40. This reduces to <1% of cases at age 68 (vs 81% with PCE) (**Supp. Fig 2)**. At age 40, MSGene had substantially greater sensitivity for lifetime CAD events compared to PCE (event reclassification 58.2%, 95% CI 58.1–58.3%), at the cost of moderate inappropriate up-classification of lifetime non-events (non-event reclassification –37.3%, 95% CI 37.2–37.4%). At age 70, MSGene had substantially greater specificity compared to PCE (non-event reclassification 32.1%, 95% CI 31.9–32.1%), at the cost of some inappropriate down classification of events (event reclassification –12.5%, 95% CI –12.4 to –12.6%). Overall, reclassification was consistently favorable (median NRI 0.12) over 40 years of consideration. Furthermore, 9.7% (95% CI 9.6–9.8%) of individuals in the top 20% of genetic risk are identified to have greater than 10% MSGene predicted lifetime risk, while only 3.1% (95% CI 2.9–3.2%) of those in the bottom 20% of genetic risk achieve this level of risk (**Supp. Fig 2)**.

### Improved calibration when compared to 30-year risk score

MSGene had improved results when compared to FRS30RC. We compared the average predicted risk by sex and genomic risk strata with empirical overall incidence rates. In healthy individuals, the RMSE of MSGene is 1.06% (1.04% males, 1.09% females, SEM 0.06) while FRS30RC is 10.9% (12.1% males, 10.1% females, SEM 0.07, **Supp. Fig. 3)**. FRS30RC increases monotonically across the lifespan. When restricting the analysis to ages 40 and 50 for whom 30 years of follow-up is available, the RMSE is 0.98% with MSGene when compared to 5.68% for FRS30RC. We further compute the RMSE starting from additional single-risk factor phenotype states (hypertension, hyperlipidemia, and diabetes) across a grid of covariate choices (**Supp. Table 1)**.

### Dynamic effects of 10-year, 30-year and remaining lifetime risk

MSGene allows for the estimation of survival curves for an individual starting from a given age, and for updated remaining lifetime curves asked over a range of ages. We compute the remaining lifetime risk when compared with FRS30RC, as recalibrated for our population.^28^ First, we depict the predicted survival curve for individuals of six different genetic and sex strata starting in health at age 40. Under this traditional analysis, CAD-free survival is projected to decline monotonically as a function of sex and genetic risk to 96.8% (95% CI 96.78–96.82) for a female in the lowest genetic strata and to 81.26% (95% CI 81.24–81.28) for a male in the highest genetic strata. However, a remaining lifetime risk curve reveals opposite behavior: for example, a high genetic-risk male has a 22.9% (95% CI 22.7–23.1%) risk without treatment at age 40, but the same high-risk male has only a 10.21% (95% CI 10.20–10.22%) risk of developing CAD if he remains CAD-free at age 70. This contradicts the 10-year risk prediction, in which 10-year risk rises from 2.84% at age 40 to 10.21% at age 70 (**Fig. 3, Supp. Tables 2– 17**). We compare this to FRS30RC projections^26^ and note that while remaining lifetime risk declines with age, the extended fixed-window (FRS30RC) approach shows monotonically increasing risk across genetic strata. In our cohort the FRS30RC risk for a high genetic-risk male rises from 13.4% at age 40 to 33.0% (**Fig. 3**) at age 70 using the recalibrated measure. When applying trial-estimated statin benefit via introducing a trial-estimated relative risk reduction to each annual transition probability^29^ (**Methods, Eqn. 2)** under MSGene lifetime projections, predicted absolute risk under treatment for the same high-genetic-risk male at age 40 improves from 22.86% (95% CI 22.85–22.87%) to 18.70% (95% CI 18.69–18.71%) over the 40-year span. This is compared to a smaller decline from 10.21% (95% CI 10.19–10.22%) to 8.25% (95% CI 8.24–8.26%) at age 70.

**Figure 3.**
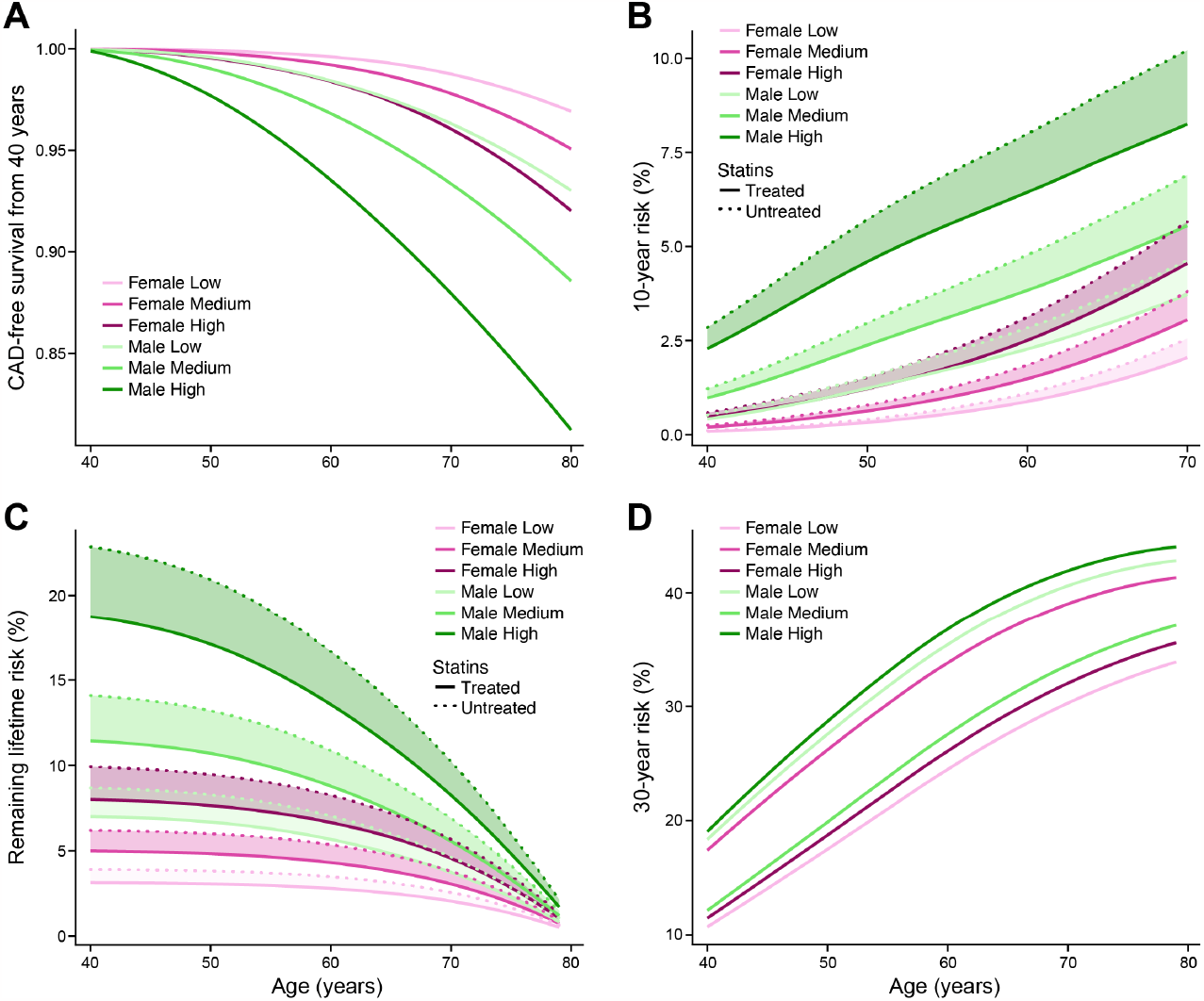
Survival, 10-year and lifetime risk curves. In **A**., we demonstrate the singular projected survival curve by MSGene for an individual at age 40 of low, medium or high genomic risk. In **B**. we demonstrate the MSGene predicted 10-year risk for individuals at each age along the x-axis, showing that, in general, for fixed window approaches, 10-year risk is monotonically increasing. In **C**, we demonstrate the MSGene predicted lifetime risk curve for individuals at each age featured along the x-axis under an untreated (dashed) or treated (solid) strategy. The conditional remaining lifetime risk declines with age, from 24% for a high genomic risk individual in our cohort to <5% for an individual at the same risk level by age 70. In **D**, using the FRS30RC equation, like 10-year risk and unlike the remaining lifetime risk approach, 30-year risk calculation is monotonically increasing, from 13.4 (13.2–13.6%) at age 40 to 32.9% at age 70 for an individual of the highest genomic risk. **FRS30RC:** Framingham 30 year recalibrated.

### Dynamic prediction: Model assessment

An updated lifetime prediction, conditional on a patient’s current state, can be made per year, using age-specific coefficients. We use these updated predictions as covariates in a time-dependent Cox model to evaluate the performance of our model on predicting time to event (**Methods**). We first consider the age distribution at which an individual first exceeded a lifetime risk threshold of 10% using MSGene or FRS30RC, or using a PCE-derived 10-year risk threshold of >5%. Using MSGene to assess lifetime risk, 44.8% percent of individuals exceed this threshold at age 40 while 38.9% never do. With FRS30RC, 44.1% exceed this threshold at age 40, but virtually all (99.8%) exceed this threshold by age 80. Using the first age exceeded under each model as a time-dependent predictor of CAD status, we find that MSGene improves model concordance by 21% (C-index 0.73 vs 0.52, *p* < 2 × 10^−16^) and of the 10-year index by 17.4% (C-index 0.55, *p* < 2 × 10^−16^) (**Fig. 4a-d)**.

**Figure 4:**
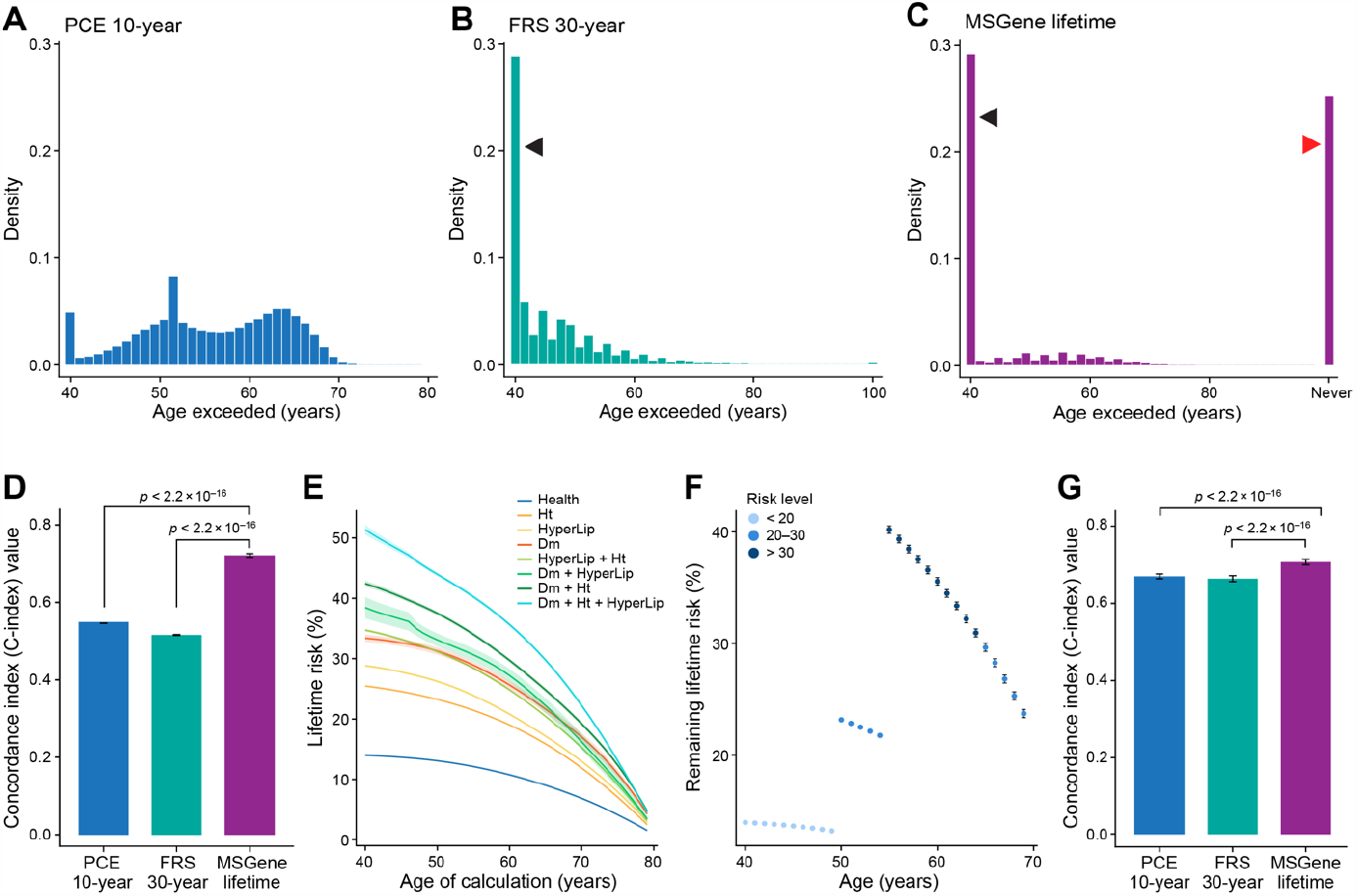
Time-dependent threshold analysis. We consider the distribution of the first age at which an individual exceeds the PCE-derived 10-year threshold of 5% (**A**), or lifetime threshold or 10% using FRS30RC (**B**) or the MSGene lifetime prediction (**C**). We then use this age as a time-dependent predictor of time-to-event in a time-dependent Cox PH (**Supp. methods)** in which an individual’s time followed is stratified by start time and periods in which a threshold is passed, and final censoring time with an indicator variable demarcating whether or not each threshold has been surpassed. We left censor these intervals at age of enrollment conservatively to exclude time protected from death. We report Harrell’s C-index (*p* < 2 × 10^−16^) for discrimination on how well a model predicts events that tend to occur earlier versus later. Left-facing indicate individuals who surpass the threshold at first prediction, and right-facing arrow indicates individuals who never surpass a threshold for a given metric. FRS30RC is shown here with C-index 0.52 (original FRS30 C-index 0.50) vs. MSGene 0.72, *p* < 2 × 10^−16^) (**D**). We compute the lifetime prediction at each age under one of eight potential risk starting states, with bootstrapped confidence intervals for a sample individual (**E**). Using the electronic health record, we extract state position for each individual per year. We then use MSGene to compute predicted risk for each individual at each state in time, displayed here for a sample individuals (**F**). We use these as predictors in a time-dependent Cox model in which we expand the data set into non-overlapping intervals for each individual (**Supp. methods; Supp. Fig. 17)** and conservatively left censor before enrollment to avoid time protected from death. We evaluate the concordance when compared to FRS30RC and PCE-derived 10-year, *p* < 2.2 × 10^−16^ (**G**). **FRS30RC:** Framingham 30-year recalibrated, **PCE**: pooled cohort equations, **Cox PH**: Cox proportional hazards model

We then use the yearly time- and state-varying predictions as predictors in a time-dependent Cox proportional hazard model in which one’s score is recorded annually in non-overlapping intervals and estimate the concordance of this model. The concordance of this time-dependent model using dynamic MSGene predictions exceeds that of the updated FRS30RC predictions by 0.71 vs 0.66, *p* < 2 × 10^−16^ (**Fig. 4e-g)**. We repeat these results using the subset with general practice (GP) records alone for both training (80%) and testing (20%) and the results hold for both the thresholding analysis (C-index 0.71 vs 0.53, *p* < 2 × 10^−16^) and continuous time-dependent analysis (C-index 0.73 vs 0.67, *p* < 2 × 10^−16^, **Supp. Fig. 4-5**).

### Estimated benefit

Our model incorporates the estimated benefit of a treatment strategy, assessed conditional on starting age and risk status. Using a randomized clinical trial (RCT)-imputed annual risk reduction of 20% for statins on statin-free individuals,^30,31^ we observe an inverse relationship between predicted 10-year risk and expected benefit. An individual with the highest genetic risk at age 40 has a predicted 10-year risk (4.2%, SD 0.01) roughly equivalent to the lowest genetic-risk individual at age 70 (3.9%, SD 0.01), but an expected lifetime absolute risk reduction of 5% (SD 0.01) at age 40 versus only 0.8% (SD 5 × 10^−2^) at age 70 (**Fig. 5**). When we consider the distribution of all starting states, we see that the mean absolute risk reduction is the greatest for younger individuals (4.6–7.2%; SD 0.01) across risk states at age 40, to a mean absolute risk reduction of 0.3–3.5% (SD 0.01) at age 79.

**Figure 5:**
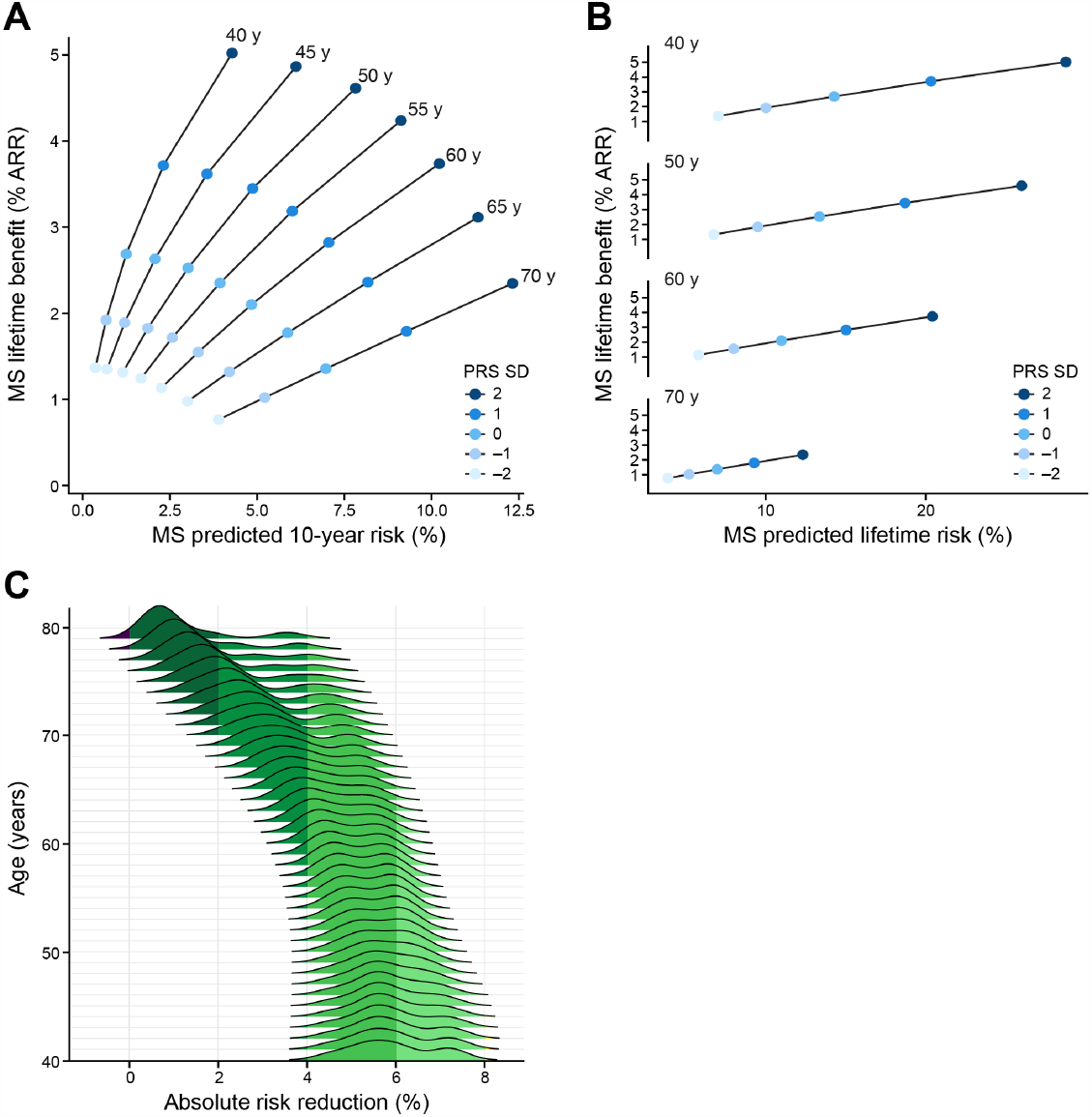
Absolute risk reduction: Short-term and lifetime risk. We display the relationship between remaining lifetime and 10-year risk. Each ray represents an age group, in which individuals are parameterized by their short-(10-year) and long-term (lifetime) risk, and colored by genomic risk in SD from mean. We display the lifetime absolute risk reduction as computed in Equation RR and stratified by age rays, and colored by genetic risk. (**A**) For an individual at the top genetic risk at age 40, MSGene predicted 10-year risk is roughly equivalent to an individual at the lowest genetic risk at age 70 (3.8% vs 4.2%, SE 0.01). However, the MSGene projected lifetime benefit is directly proportional to lifetime risk (**B**), and more than twice that of a high risk individual at age 70 (5.0 vs 2.3%, SEM 0.02). (**C**) Marginalized across starting states and covariate profiles, we project absolute risk difference (%) under a treated and untreated setting. At age 40, this ranges from a median of 5.8% (SD 0.01) to 0.8% (SD 0.01) at age 79. **SEM:** standard error of mean, **RR:** relative risk, **SD:** CAD-PRS SD.

### Improvement in discrimination over the cumulative horizon

When considering only the presence or absence of disease over observed time without regard to timing, the AUC–ROC of a model comparing the prediction of cumulative occurrence using updated MSGene lifetime score shows greater performance than that of either FRS30 or FRSRC early in the life course (**Supp. Fig. 6**) (0.69 vs. 0.65, *p* < 2 × 10^−16^ at age 40) and also based on precision-recall (0.20 vs 0.16 at age 40, *p* < 0.01). Both metrics exceeded the estimation of lifetime risk using genetics as a predictor alone. In general, when comparing individuals captured by MSGene but not by FRS30RC, MSGene identified more women and individuals at higher genetic risk. With time, these differences were more profound (**Supp. Fig. 7**).

### External validation

We then performed external validation of MSGene in the FOS cohort, using first measurements to ensure optimal follow-up duration. FOS is a community-based cohort recruited in 1971 with a median 39 years of follow-up [IQR 38–40], median age of enrollment 35 years [IQR 28–44] (**Supp. Fig. 8**). MSGene again had favorable discrimination (age 40: 0.75 [95% CI 0.69–0.82] vs. 0.73 [95% CI 0.66–0.80]; age 55: 0.63 [95% CI 0.42–0.84] vs. 0.53 [95% CI 0.29–0.76]) and calibration (RMSE 8.4% vs. 11.3%, *p* < 2 × 10^−16^) when compared to FRS30 (**Supp. Fig. 9**).

## Discussion

This study introduces a novel method called MSGene, which aims to assess the risk of developing CAD and other health states over the lifespan. Our dynamic lifetime risk predictions improve considerably calibration and discrimination and improve the identification of younger individuals at high risk without overestimating risk in older adults, compared to previous models. Our projected benefit analysis shows large reduction in preventable CAD events if statin therapy is guided by MSGene.

The technique utilizes generalized linear models (GLMs) to compute the transition probabilities between different states (e.g., from a healthy state or risk factor to CAD, death, or intermediate risk) for every age over the observed life span. The novelty derives from four features: 1) the provision of unique age-dependent models via GLMs that allow the relationship of each covariate on the outcome to vary freely with time; 2) the calculation of risk conditional on time-dependent states; 3) the assessment of a multistate model via time-dependent Cox modeling; and 4) the unique use of the UKB EHR as a comprehensive longitudinal data resource. The study follows individuals from adulthood through their enrollment in the linked health record. By incorporating age and time dependence, this method provides annual risk estimates that include the entire lifespan.

Over a lifetime horizon, the dynamic change in risk makes accurate lifetime risk estimations challenging.^4,7,11^ However, leveraging genetics **in addition to multi-state modeling**, MSGene enhances lifetime risk predictions, effectively identifying individuals previously deemed low-risk MSGene enhances lifetime risk predictions, effectively identifying individuals previously deemed low-risk. The model’s age-dependent features, producing age-sensitive coefficients, negate the need to rely on fixed parametric interactions between each covariate and time, a prevalent limitation in traditional models.^6^ We show that using updated estimates conditional on the dynamic state of an individual improves *time-to-event* prediction overall.

Through the incorporation of treatment, we show that those individuals with the greatest and least expected absolute risk reduction from statin therapy actually have a similar 10-year risk. However, this short-term focus is what current clinical methods rely upon.^7^ Presented effects are conservative as statin effects may magnify with duration and on CAD PRS background.^19,32–34^

Our approach facilitates accurate event prediction both for undercaptured young individuals and also lower-risk older individuals who might otherwise be included in a fixed-window approach that extends the time horizon: our median global net reclassification when compared with a 10-year approach is 12.2% [IQR 5.5–18.6%] over 40 years. This in part explains the improvement in overall time-dependent performance when incorporated into a time-to-event framework. Using a time-dependent evaluation, the distribution of the first age at which a lifetime threshold is exceeded demonstrates that MSGene optimally identifies at-risk individuals without indiscriminately calling all patients ‘at-risk’. However, future work is warranted to determine optimal thresholds of lifetime risk to maximize potential benefits among high-risk younger individuals while reducing unnecessary costs and harms to low-risk older individuals.

One of the strengths of our method is the access to a significant history of electronic health records that allow us to derive estimates informed by a greater group of patients throughout the life course. Existing scores^26,35^ imply that the levels of covariates will stay fixed over the life course or require recalculation, which ignores the information within transitions through the life course. Here, our longitudinal outlooks ability allows for individuals to be followed over a lifetime and quickly estimates what their updated risk trajectory would look like under an alternative profile.

Estimation of remaining lifetime risk is conducted using age-specific predictions informed only by individuals in the at-risk set at a given age, thus making this a true lifetime estimate. In our work, we choose a conservatively estimated age of 80 as the maximum lifetime age given the density of age estimation with our set. This estimation is possible under the assumption that risk trajectory is similar across shifting windows of age at risk but falls apart with strong calendar time trends. Given that our cohort was required to be between 40 and 70 years old in 2006, we reduced the variation in calendar effects.^5,36^

When combined with genetic information, an emphasis on dynamically updated lifetime risk projections can uncover latent risks in seemingly healthy individuals. Determining an appropriate lifetime risk threshold is a laudable goal.^2,7^ Indeed, current guidelines^12,36^ note that genetic risk scores can identify individuals at birth with a high propensity to develop disease, but few approaches have coupled this information with realized risk stages dynamically. As age increases, short-term risk increases, and the remaining lifetime risk is reduced, meaning that a metric focusing on short-term risk will preferentially focus on disease in older individuals, thwarting the efforts of true prevention. It is not enough to increase the lifetime threshold to account for younger individuals as proposed in European Society of Cardiology guidelines; additional years add additional uncertainty, and thus, having tools capable of dynamically incorporating new information over the life course in combination with more comprehensive time assessments is critical to moving prevention forward. We provide an application for individuals to assess risk in real-time for patients and clinicians **(Supp. Fig. 10**; **https://surbut.shinyapps.io/risk/)**

In this study, we use a composite of phenotypic codes to define our risk factor states. One of the challenges of developing a lifetime assessment tool surrounds the availability of continuously updated laboratory data. Using EHR data, an unbiased ascertainment of updated biometric variables at uniform intervals is challenging. We added baseline continuous laboratory data from the age of enrollment to our grid search, and this added little to our model (**Supp. Fig. 11)**.

A second limitation surrounds the heterogeneity of phenotyping. We define hyperlipidemia and hypertension according to validated diagnostic codes.^37^ However, there exists heterogeneity in the severity and duration of these conditions. The potential benefit of adding additional states must be balanced with the uncertainty imposed and the reduction in sample size caused by dispersion across grades of each condition. Our model resolves the loss in underlying latent risk that is often erroneously captured in EHR data when an individual’s nominal laboratory value falls secondary to medication use.

One of the advantages of heterogenous data collection is a wealth of available phenotyping modalities: the UKBB has access through linkages to routinely available national health systems enhanced by self-report and previous records.^38^ Although not all individuals included had GP data, we demonstrate that the age and prevalence of conditions is homogenous between individuals in the GP subset and otherwise (**Supp. Figs. 1**) and that analysis on this subset alone results in similar model discrimination.

Third, the generalizability of our findings may be impacted by study design and sample specificity. The UK Biobank included healthier and less socioeconomically deprived individuals who were predominantly White Europeans living in the United Kingdom.^39^ Furthermore, given that the minimum age for genotyping was 40 years old, we began our inference for risk modeling at age 40, provided they were captured in the EHR before then. Although individuals who reached age 40 prior to enrollment were appropriately at risk for the primary CAD outcome given their capture in the longitudinal EHR, they were protected from death until the time of enrollment, which may affect estimates related to the competing risk of death. For time-dependent evaluation of our prediction, we conservatively left-censored at age of enrollment to eliminate years protected from death and found that the improvements in discrimination over FRS30RC remained unchanged. We note consistent performance in external validation in the FOS cohort, where all death and CAD events occurred exclusively after enrollment. Finally, our dynamic logistic regression approach can readily be adapted to any population with minimal computational resources, and we provide code to do so.

Leveraging a unique resource of genetic and longitudinal clinical data spanning over 80 years in nearly 500,000 participants of the UK Biobank prospective cohort study, we develop MSGene, a multistate model for dynamic transitions throughout the life course to estimate lifetime risk of CAD. MSGene is well-calibrated and discriminates early and late events both in the UK Biobank and an external validation sample. We anticipate that by providing interpretable and dynamic estimates of CAD lifetime risk, MSGene may inform future therapeutic decisions to enable more efficient and effective CAD prevention throughout the life course.

## Data Availability

All code for running the MSGene model is available at https://github.com/surbut/MSGene. Vignettes for running the analyses are available at https://surbut.github.io/MSGene/vignette.html and https://surbut.github.io/MSGene/usingMarginal.html. Shiny app for calculating interval risk is available at https://surbut.shinyapps.io/risk/. UK Biobank data is available upon application through the UKB Showcase https://www.ukbiobank.ac.uk. Framingham Offspring Data is available through dbGap access by investigator application.

## Acknowledgments

We would like to acknowledge Leslie Gaffney of the MIT-Broad Communications Lab for her invaluable graphics and copyediting advice.

## Methods

### Data source

The UK Biobank (UKB) is a prospective UK population-based study that enrolled approximately half a million adults aged 40–70 between 2006 and 2010 designed to investigate the genetic and lifestyle determinants for a wide range of diseases. Participants underwent genome-wide genotyping, with linkage to longitudinal hospitalization, primary care (GP), and self-report data dating back to 1940 (**Fig. 2; Supp. Figs. 12-13**).^37^ Using the ***ukbpheno*** package (version 1.0),^37^ we assembled detailed longitudinal data from the various sources documenting events from 1940 until December 2021 for 481,927 individuals after excluding 20,534 who lacked quality control genotyping or risk factor information **(Fig. 2; Supp. Fig. 12-14)**. At the time of analysis, linkage to the United Kingdom General Practice (GP) Registry was available for a subset of 221,351 individuals. This assembly across data-sources generated phenotypes for hypertension (Htn), diabetes mellitus (DM) (type 1 or 2), hyperlipidemia (Hld), or coronary artery disease (CAD) based on validated collections of hospitalization (HESIN), diagnostic, operation, general practice (GP) clinical and script as well as death information.^37^ We found high overlap between these phenotypes and our own lab’s previously generated HESIN-restricted phenotypes^32,40^ (**Supp. Fig. 14**). These phenotypes subsequently became the risk factor states in our model. Informed consent was obtained from all participants, and secondary data analyses were approved by the Mass General Brigham Institutional Review Board 2021P002228. Secondary data analysis of UKB was performed under application number 7089.

Because of the longitudinal nature of this cohort, every individual is observed at first encounter with the electronic health record (EHR) in early adulthood (median age 24.2 years). We selected UKB participants free of CAD at age 40 and followed until the occurrence of CAD, death, or loss to follow-up (median follow-up 29.9 years). We categorize individuals by their condition at entry into our cohort at age 40 provided they have been observed in the EHR (**Fig. 2**). We then re-evaluate at each age the risk set as those individuals who have 1) been observed and 2) have not been censored for a given phenotype. We demonstrate the diversity of data sources and the corresponding availability of each data source over time for all considered phenotypes (**Supp. Fig. 13)**. In general, our model allows for the progression from CAD to death, but we report here the risk of progression to CAD on CAD-frr individuals at baseline.

### Polygenic risk

An additional novelty of our model is the incorporation of the dynamic effects of genetics over time. We use CAD polygenic risk score (PRS) as released through the UKB resource^41^ and compute on individuals with adequate genotype information after quality control and after controlling for the principal component axes obtained from the common genotype data in the 1000 Genomes reference data set using standard methods^41^. Data supporting these scores were entirely from external GWAS data (the Standard PRS set) as conducted by Genomics PLC (Oxford, UK) under UKB project 9659.^41^ We demonstrate that the distribution of PRS is similar across entry age (**Supplementary Figure 15**).

### Statistical analysis Detailed Equations

Let *π*_*jkia*_ represent the annual transition probability from state **j** to state **k** for individual **i** during year **a**. We let the states **j** and **k** represent time-dependent phenotypes ascertained from the electronic health record such that every individual is in the at-risk ‘healthy’ set until first censoring. For p-covariates for a given individual transitioning from state **j** to **k** we refer to the following equation. ‘From’ states **J** include Health; single risk factor states: Hypertension (Ht), Hyperlipidemia (Hld), Diabetes Mellitus Type 1 and Type 2 (DM), double risk factor states: Ht & Hld, Ht & Dm, Dm & Hld; Triple risk factor states: Dm & Hld & Ht; and Coronary Artery Disease (CAD). States **K** include all of the ‘From’ states and Death. For our purposes, we report the progression to CAD or death from any of the starting states included in J.

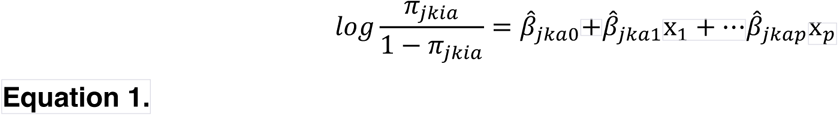

Taking the inverse logit of the estimate returns the absolute risk for any individual **i** is a function of the age-specific coefficients and his **p** covariates, such that the annual risk estimate from state **j** to state **k** satisfies:

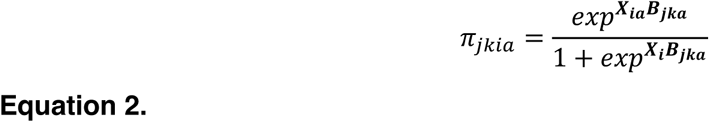

Here we let ***X*** represent the N x P matrix of individuals and covariate profiles at a given age and ***β*** represents the P x 1 vector of age and state-state specific smoothed coefficients.

In equation **2**, state **j** represents the ‘from’ state and state **k** represents the ‘to’ state. To account for censoring, an individual exits the ‘at risk’ group for transition inference when they are lost to follow-up. We use a one-year interval over which to discretize age intervals and independently estimate the *π*_*jkia*_ age-dependent-state to state transitions. We use a limited number of time-fixed covariates: that is sex and polygenic risk score (PRS) and estimate time-dependent effects. We assess current smoker at enrollment in the UK Biobank and use as a time fixed effect for model estimation – that is, individuals reporting ‘current smoker’ at enrollment in the UKBB are considered as smokers in each age-specific logistic regression. For inference of time-dependent covariates, we treat both anti-hypertensive and statin use as individual time-dependent covariate which is reevaluated at each year of model estimation using prescription data from the UKB.^42^ Our final prediction model allows for continuous updates of smoking and medication usage in estimating age-specific transition probabilities. We use 80% of our data as training and 20% as testing (**Fig. 2**) for internal cross-validation and to optimize model fit. Accordingly, this divides our data into a training set for model fitting using 384,510 samples and a testing data set of 79,117 unique individuals.

### Predicted Interval Risk

Predicted risk over a given time interval for a given individual **i** of progressing to state **k** from state **j** over any Y-year period ranging from age **A1** to **A2** is where **a** indexes the current age:

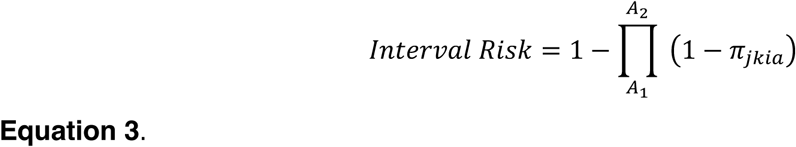

Accordingly, risk for an individual **i** of progressing to state **k** from state **j** where **L** is the maximum age of life and a is the currently observed age. For our purposes, we choose **L** = 80 in line with the available data by age in the UK Biobank.

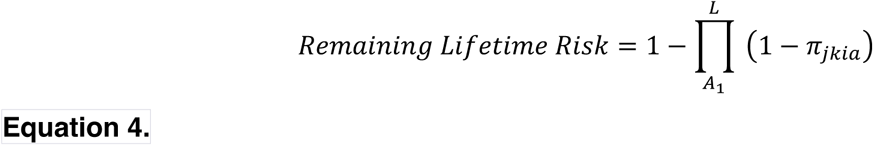

The remaining lifetime risk can be modified to account for treatments by applying a constant relative risk reduction to the age-specific transition probabilities in expression 4. Then the interval risk under treatment can be calculated using the per-year risk reduction **RR** of progressing to state k from state j over an interval from age **A1** to **A2** is:

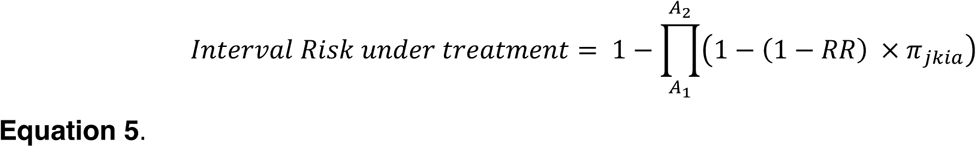

For the purposes of this manuscript, we are interested in state k = CAD. We impute the relative risk reduction of 0.20 from 24 trials of statin therapy.^29^ Within our model, we constrain each individual’s predicted probabilities across states per year to sum to one such that for each age **a**, the probability of staying within the given state is the complement of the sum of transitions over K to the alternative states:

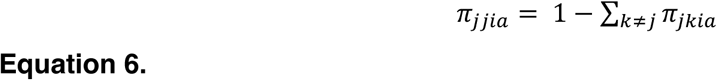

It is somewhat arbitrary to choose j as the “to” state whose probability is determined as the complement of the others. We choose j because it is mostly above 50% and the constraint in 6 will guarantee that for a given age the probabilities for an individual of a particular covariate profile sum to 1. The alternative of fitting a polytomous regression is computationally much more demanding and gives approximately the same answer.

### Flexible Smoothing Across Ages

We extract the unsmoothed coefficients 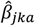 for each age and state transition from the logistic regressions in (2). To borrow information across ages, we fit a smoothed locally estimated polynomial regression in which for each state to state transition and each covariate, we fit a locally estimated weighted regression^27,43^ (LOESS) (**Supplemental Figure 16**). The loess weights are proportional to the product of the inverse variance of each estimated coefficient and the tricube distance function of nearby ages to smooth adjacent ages more closely together proportional to the cube of their distance d from the age in question:

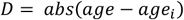

We consider the neighboring unsmoothed coefficients as those within an adjusted window length, and if the age in question is within 5 years of the minimum or maximum age, we extend the adjusted window by 5 years.

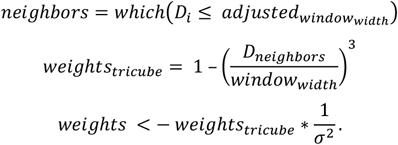

We then use weighted least square regression to adjust the coefficient as the weighted sum of neighboring coefficients where the design matrix X is the ‘N’ neighbor’ by degree +1 matrix X and y is the N x 1 vector of unsmoothed coefficients.

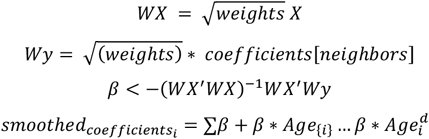

A vignette showing this process on a sample calculation is shown here https://surbut.github.io/MSGene/vignette.html. Furthermore, flexible window choices and polynomial degrees can be found here: https://surbut.shinyapps.io/testapp/. All analyses were performed with R (version 4.3.1) and our software is written as R code with implementation and vignettes at https://github.com/surbut/MSGene.

#### Standard Error of Projection

We bootstrap our training data 50 times and extract the corresponding means and standard errors of each projection across bootstrapping iterations. We compute the remaining lifetime risk setting the maximum age considered as 80 according to the density of observations in our training data, and impute a relative risk (RR) of CAD from statins of 0.20^30,44,45^; notably, the RR may be larger for some groups, such as those with elevated CAD PRS^32,46^, and for longer periods of time and thus this reflects a conservative estimate^47^. We apply this benefit only to individuals *not* previously on statins.

For the RMSE, we report the standard error of the mean across strata. For proportions, we report the standard error of the sample proportion as 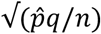 where 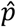 represents the sample proportion.

#### Precision and Discrimination analysis

For each age, we compare the average predicted score by genomic (<20%, 20–80%, and >80%) and sex strata, and report the root mean squared error (RMSE) as the difference in the average empirical and predicted cumulative incidence rate for each PRS and sex group as detailed in the Supplementary Methods.

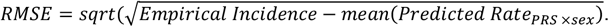

For the area under the receiver operator curve (AUC-ROC) and precision-recall analysis, we compute the area under each curve using each score as a predictor of cumulative case or control status computed using values for individuals at each year plotted.

#### States and competing risk

The unique nature of our multistate model features eight mutually exclusive states and restricts one-year transitions as follows (**Fig. 1)**, with death as the final absorbing state from which one cannot exit. At any age across the life course, cumulative one-step transitions can be assessed (**Fig. 1**). Possible transitions are as follows:

1. Health to a single risk factor (Htn, Hld, Dm), CAD or death;
2. Single risk factor to corresponding double risk factor, CAD or death;
3. Double risk factor to triple risk factor, CAD or death;
4. Triple risk factor to CAD or death;
5. CAD to death.

##### Predictions with age as the time scale

Our model inferences are made per-year using the individuals who are in a particular risk state at a given age (**Fig. 2, Supp. Fig. 12**). Predictions can, therefore, be made over a requested time interval using the product of age-specific risks for which coefficients were estimated from individuals who were in the at-risk subset during a given period. While enrollment in the UK Biobank required that an individual be alive at age 40 to enroll for genotyping, it did not require that the individual be risk factor-free, and therefore we use this information to assign individuals into risk categories for inference from age 40 onward. We exclude individuals with CAD at baseline from our predictions.

##### Comparison to 10-year PCE and 30-year Framingham CAD risks

For comparison of time-dependent 10-year risk, we use the 2018 PCE with baseline covariates (total cholesterol, HDL-cholesterol and systolic blood pressure, current smoking) obtained from UKB enrollment data and update each prediction^26^ with time-varying age, diabetes, and medication use according to available records. This technique was used in the Framingham 30-year risk development to validate new longer window estimates in which age was iteratively updated with all other risk factors at their baseline values.^26^

For comparison of calibration to 30-year risk, we used the 2009 complete (non-BMI) Framingham 30-year equation (FRS30) and update each prediction^26^ with time-varying age, diabetes, and anti-hypertensive use according to available records, consistent with detailed formulae within the FRS30. Given the differing populations, we recalibrated^48^ according to the mean levels of each covariate and baseline hazard in the UKB sample (FRS30RC). For fair comparison, we report our results against FRS30RC given its improved calibration in our cohort (**Supp. Fig. 17**). Precision and discrimination analysis described as follows. We compute and display the predicted 30-year risk for individuals from ages 40–70 according to this model.

##### Time-dependent model assessment

We first use the age and state-specific predicted risk scores for each individual - which arise from our MSGene system of smoothed logistic regressions - as covariates in a time-dependent Cox model, in which an individual is featured in non-overlapping intervals with their respective score and event status. In the evaluation stage, we conservatively left censor individuals until enrollment. We also calculate the minimum age at which an individual would exceed pre-specified risk thresholds for MSGene, PCE, and FRS30. We divide every individual’s observed trajectory into non-overlapping intervals, indicating when one or all thresholds are achieved and when an event occurs. For example, if an individual is observed from ages 40-70 and exceeds one risk score at age 45 and the other at age 52 and has an event at age 68, his period of study will be divided into 4 intervals: the period from age 40 to 44 in which he exceeds the threshold with neither score, the period from 45-51 in which he exceeds the threshold only with score 1, the period from 52 to 67 in which exceeds with both scores, and the period from 68 to 80 in which he has had an event and exceeded in both score. We left censor in this analysis at age of enrollment. We fit independent time-dependent Cox models^49^ to this expanded data set, and again conservatively left censor until enrollment. For both analyses, we report the concordance index (Harrell’s-C) with confidence intervals derived from bootstrapping iterations.^50^

##### Internal and external model assessment

We internally assess the calibration (RMSE) (**Supp. Table 1)** of models using a finite number of covariates for eight state-specific transitions built on a training set and independently assess on our testing set. External validation was performed by comparing the model fits estimated in the UKB with 10-year and lifetime risk estimates from young adults in the Framingham Heart Study Offspring cohort (FOS)^51^ (**Supp. Fig. 8)** for whom genetic data are available. This is a community-based Northeastern United States cohort that was recruited in 1971, median age [IQR] 33.0 years [27.0, 41.0] and followed through 2013. Clinical data and incident disease for 3836 participants, and genetic data for a subset (2611), were available through the database of Genotypes and Phenotypes (dbGaP; accession phs000007.v33.p14). We compare these with the PCE and FRS30 (original score, calibrated for this population) estimates calculated at Exam 1 and compute the RMSE and AUC over the 30-year follow-up period. Informed consent was obtained from all participants, and secondary data analyses of dbGAP based FOS and UKB were approved by the Mass General Brigham Institutional Review Board applications 2016P002395 and 2021P002228.

### Calculating Net Reclassification

For net reclassification indices, at each age of consideration, we defined *NRIevent* as the net proportion of cases correctly reclassified by MSGene Lifetime (MSGeneLT >10%) as compared to a ten-year PCE:

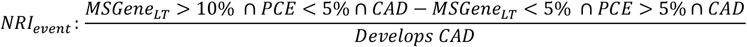

We defined *NRInon-event* as the net proportion of controls correctly reclassified by MSGene lifetime risk <10%:

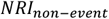

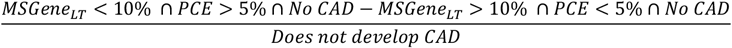

### Marginal Calculation

We also allow, for the absorbing states of CAD and death, the possibility of computing the probability of progressing through any out (‘marginal’) to CAD. The calculation of progressing to state K from state J through any path over N years is the product of N transition matrices **T** in which the **j**,**k** element for matrix **Tia** is the probability of progressing from state **j** to **k** at age **a** for individual of covariate profile **i**:

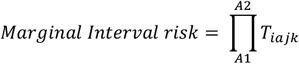

For every individual, we constrain the row sums to sum to 1 so that the marginal probability across states cannot exceed 1. For absorbing states, the k,k probability is 1. This vignette is available at https://surbut.github.io/MSGene/usingMarginal.html.

