## Supplementary figs and table 1 for "MSGene: Derivation and validation of a multistate model for lifetime risk of coronary artery disease using genetic risk and the electronic health record"

**Urbut et al.**

1. Supplemental Tables and supporting information (PDF file format)
2. Supplemental Figures and Figure Legends (PDF file format)
3. Additional Supplementary Materials

### Supplementary Tables

Supplementary Table 1

Supplementary Tables 2-16 in the Excel document

| | Health | | Hypertension | | Diabetes Mellitus | | Hyperlipidemia | |
|---|---|---|---|---|---|---|---|---|
| **RMSE (SD)** | MSGene Ten | MSGene Life | MSGene Ten | MSGene Life | MSGene Ten | MSGene Life | MSGene Ten | MSGene Life |
| Sex + PRS | 0.59 (0.03) | 2.24 (0.06) | 1.47 (0.07) | 6.44 (0.23) | 4.02 (0.18) | 8.99 (0.3) | 3.46 (0.17) | 9.93 (0.36) |
| Sex + PRS+Smoking (RMSE (%)) | 0.61 (0.03) | 2.34 (0.07) | 1.48 (0.07) | 6.53 (0.23) | 4.10 (0.19) | 9.40 (0.31) | 3.49 (0.17) | 10.05 (0.36) |
| Sex + PRS+Smoking+ Antihypertensive | 0.68 (0.03) | 1.34 (0.05) | 1.46 (0.07) | 6.33 (0.23) | 3.96 (0.19) | 8.51 (0.30) | 3.31 (0.16) | 8.58 (0.31) |
| Sex + PRS+ Smoking+ Statin | 0.74 (0.04) | 1.13 (0.03) | 1.36 (0.07) | 5.10 (0.16) | 3.93 (0.18) | 9.15 (0.31) | 3.49 (0.17) | 9.83 (0.35) |
| Sex + PRS+ Smoking+ Antihypertensive + Statin | 0.86 (0.05) | 1.06 (0.04) | 1.36 (0.07) | 5.4 (0.17) | 3.93 (0.19) | 8.65 (0.30) | 3.33 (0.16) | 9.01 (0.32) |
| Pooled Cohort Equation | 6.12 (0.32) | | 6.74 (0.37) | | 10.93 (0.54) | | 7.03 (0.34) | |
| Pooled Cohort Equation (restricted to individuals at enrollment) | 6.08 (0.31) | | 7.10 (0.36) | | 7.25 (0.37) | | 7.10 (0.35) | |
| FRS30 Year[15] | | 33.60 (0.75) | | 37.45 (0.83) | | 42.67 (0.82) | | 35.91 (0.87) |
| FRS30 Year Recalibrated | | 10.91 (0.26) | | 12.25 (0.34) | | 16.76 (0.46) | | 12.58 (0.4) |

**Supplementary Table 1: RMSE (%) of limited grid search for model fit**

Above, we demonstrate the RMSE of each model using a set of covariates comparable to existing risk stratification algorithms for individuals for prediction over ages 40-70. Each RMSE is averaged over a set of sex, genetic and age strata, as described in text. We provide SEM for RMSE across strata. We compare the Pooled Cohort Equation (PCE) ten-year risk for individuals using baseline parameters with continuously updated ages as in the original 30 year validation study[15], and to a restricted set of individuals who contribute baseline parameters at age of enrollment considered. This technique was used in the development of the initial Framingham 30-year score in 2009: namely, using baseline values of covariates and updated age to calculate risk in a model requiring these covariates. For FRS 30 year we also use the baseline values of systolic blood pressure, high-density lipoprotein and total cholesterol, with updated ages[15] and for the recalibrated calculation, we recalibrate the prediction using the mean values of covariates at baseline and the population baseline hazard as in published[16] recalibration.

**FRS30**: Framingham 30 year, **FRS30 Recalibrated**: Framingham 30 recalibrated, **SEM:** standard error of mean.

**Supplementary Figure Legends**

**Supplementary Figure 1: Summary of GP and non-GP members**
Above, we demonstrate the homogeneity of phenotyping age and proportions among individuals within and outside of the GP (general practice) cohort. We use approximately 80% (385,541) individuals in the training, and 79,119 in the testing set, of which approximately 45% represent members of the general practice primary care data.

**Supplementary Figure 2. Comparison to ten-year pooled cohort equations**
**A**. We display the proportion of cases captured using a pooled-cohort equation (PCE) threshold of 5%, a lifetime threshold of 10% as computed by MSGene, or both. At age 40, 58% of individuals who ultimately develop CAD demonstrate an MSGene lifetime threshold greater than 10% while less than 1.3% demonstrate a PCE 10-year threshold than 5% alone. **B.** The net proportion of events (NRI case) detected by a lifetime score exceeds that of a 10-year score at age 40 and the net proportion of non-events exceeds that of a 10-year measure after age 60. Median NRI over the 40-year period is 12.2% (5.4%–18.6%) **C.** High lifetime risk individuals not captured by the 10-year equation are enriched in high-genomic risk. After age 68, there are no individuals with lifetime score over 10% who lack a short term risk greater than 5%.

**PCE**: pooled cohort equations, **PRS**: polygenic risk score, **NRI**: net reclassification index.

**Supplementary Figure 3: Overall Calibration from health state.**
We display the RMSE (SEM) between predicted and realized risk for individuals starting in the healthy state by sex and genetic risk level as categorized low (<20%), mid (20-80%) and high (>80%). We also compare to the Framingham 30-year score (FRS30) and Framingham 30-year score after recalibration (FRS30RC). Here the standard errors represent the standard deviation in calibration across age, sex and genetic categories for a given score to demonstrate variability in performance across categories.

**RMSE**: Root mean squared error, **FRS**: Framingham 30-year risk score**. FRS30RC**: (recalibrated). **SEM:** Standard error of mean

**Supplementary Figure 4: Analyses using the first-age at which threshold surpassed using GP cohort alone.**
Using only the individuals in the GP cohort for testing and training, we consider the distribution of the first age at which an individual exceeds the PCE-derived ten year threshold of 5%, or lifetime threshold or 10% using FRS30RC **(B)** or the MSGene lifetime prediction **(C)**. We then use this age as a time dependent predictor of time to event in a time-dependent cox PH in which an individual's time followed is stratified by start time and periods in which a threshold is passed, and final censoring time with an indicator variable demarcating whether or not each threshold has been surpassed. We report Harrell's C-index (p <2e-16) for discrimination on how well a model predicts events that tend to occur earlier versus later. CI calculated over 100 bootstrapping intervals of expanded data set.

**FRS30RC:** Framingham 30-year recalibrated. **PCE**: Pooled Cohort equations.**GP:** General Practice cohort.

**Supplementary Figure 5: Analysis of time-to-event discrimination using the GP cohort alone.**

Using only the individuals in the GP cohort for testing and training, we use continuously updated predictions assembled combining age-specific state status information with state-specific model predictions, as in the primary analysis featured in main Figure 6. We show that the C index using MSGene updated estimates exceeds that of using the FRS30RC score ($p < 2e-16$). CI calculated over 100 bootstrapping intervals of expanded data set.

**FRS30RC:** Framingham 30-year recalibrated. **GP:** general practice cohort.

**Supplementary Figure 6: AUC-ROC**
We report the area under the receiver operating curve (ROC) predicting remaining lifetime risk using empirical data as the gold standard. We dynamically update the age along the x axis and compare to FRS30, FRS30RC, or PRS alone. We also display the precision recall curve, which accounts for class distribution changes over the life course. Here we report the ROC for the transition from health to CAD. Standard deviation represents the square root of the variance of the ROC estimate using pROC (version 1.17.4).

**FRS30**: Framingham Risk Score 30year, **FRS30RC**: Framingham Risk Score 30year Recalibrated, **AUC-ROC**: Area under the receiver operator curve; **AUC-PRC**: Area under the Precision recall curve.

**Supplementary Figure 7: Unique individuals identified.**
Comparison of individuals identified at each age by an MSGene lifetime score (using a threshold of 10%) only or by FRS30RC **(A),** PCE **(B)** or MSGene marginally. We note that after age 70, there are no individuals identified by MSGene who are not also identified by the PCE or FRS30RC metric owing to the specificity of MSGene.

**FRS30RC: Framingham 30 year recalibrated, PCE: Pooled Cohort equations.**

**Supplementary Figure 8: Framingham Offspring Cohort**
Using the Framingham Offspring cohort (FOS), we isolate individuals with genotype information available for polygenic risk scoring and use values at first measurement to compute predicted 30-year score and MSGene lifetime score. We compare with the score based on training values computed using the UKB EHR and calculate RMSE and ROC-AUC. In **(B)**, we describe the cohort over a median of 38.4 years (IQR 4.1) years of follow up. Low genomic risk connotes individuals in the lowest (<20%) of genomic risk by PRS percentile, intermediate (20-80%) PRS percentile, and high denotes >80% PRS percentile. Given the size of the cohort, we report age-specific AUC for 5-year age intervals.

**FOS:** Framingham Heart Study Offspring Cohort, **CAD:** coronary artery disease, **PRS:** Polygenic Risk score**. Pheno:** phenotyped outcomes, **RMSE**: Root Mean Squared Error, **AUC**: Area under the receiver operating curve.

**Supplementary Figure 9: External Validation**

We compute the root mean squared error (RMSE) and AUC-ROC curve for prediction for all individuals in the FOS cohort using MSGene lifetime prediction and FRS 30 in blue. Given the limited number of individuals we report across all individuals rather than by age and sex category. B) We compute the area under the ROC curve using an MSGene score for individuals starting at ages 40, 45, 50 or 55 in the FOS and compare with computed FRS30 score on 30

years of follow-up data, given that we compare with the original FRS 30-year score (calibrated on this population).
**FOS**: Framingham Offspring Cohort; **MSLife**: MSGene Lifetime evaluation; **FRS30:** Framingham 30-year score (original), **AUC=ROC:** area under receiver operator curve.

**Supplementary Figure 10: Interactive application for lifetime risk reduction**
Using our interactive application, patient's and clinicians can visualize the estimated risk trajectory based on starting CAD and covariate profile and adjust for treatment start time, changing covariate profile, and changing state. The app can be accessed at https://surbut.github.io/risk.

**Supplementary Figure 11: Model fit attempt using baseline covariates.**
We look at the estimated coefficients over 40 years of prediction for a model including baseline covariates and see that the coefficient for these values approaches after inclusion of hypertension and hyperlipidemia in a multistate approach. Given the further limitations of obtaining accurate levels of these covariates at regular intervals in an observational cohort, we choose a model that uses risk factors as opposed to individual laboratory measurements.

**CAD-PRS:** Polygenic risk score, **Anti-hypertensive use**; time-dependent antihypertensive use; **Statin Use**: time dependent statin use; **HDL-C**: HDL cholesterol: SBP: systolic blood-pressure.

**Supplementary Figure 12: Mapping the life course using EHR data**
In **A**, we demonstrate the data encountered across modalities of the UKB EHR data for a sample individual with periods of data observation from 1990 through the present who had an MI in 2013 at age 57. In **B**, for a different individual, we demonstrate the use of diagnostic code assemblies from a variety of sources including touchscreen (**TS**), self report (**f.20002),** primary care (**CTV3**) and HESIN[17] (**ICD10**) to define phenotypes of interest. This patient enters our study at first interaction with GP record in 1995 and is characterized in the hypertensive risk category. He is then later diagnosed with CAD in 2012. **C**. We show the density of first reported encounter with the primary care atlas for individuals within the UKB. Peak density between 1980-1987.
**TS**: Touchscreen; **f.20002:** Self-report, **CTV3**: primary care, **ICD10**: International Consortium on Disease. **CAD**: coronary artery disease. **EHR**: Electronic Health Record.

**Supplementary Figure 13: Availability of phenotype by data source**

Above, for the states of interest, we demonstrate the enrichment by data source for categories of codes recorded that inform our phenotyping algorithm. In general, across categories and phenotypes, diagnoses begin in 1940 and exceed 1000 diagnoses by 1980. Plots generated using the ukbpheno package Version 1.0.[18]
**Ts=**Touchscreen**, HESIN:** Hospitalization index data**, sr:** self report**, tte:** time to event**, gpclinical:** general practice clinical data**.**

**Supplementary Figure 14: Alignment of phenotypes**
Above, we demonstrate the concordance of phenotype data between the diagnoses assembled using the UKBPheno package[18] across GP and HESIN codes, and with our previously published[9,19,20] laboratory data.
**Lab:** previously published phenotypes. **UKBPheno:** using the **UKBpheno** atlas.

**Supplementary Figure 15: PRS-Distribution by Age at Enrollment in UKB**
We demonstrate the distribution of genomic risk (PRS) by age of enrollment. In general, there exists no bias between individuals who enroll at early or late ages by genomic risk quintile (*p = 0.28,* Anderson Darling for difference in distribution*)*.
**PRS:** Polygenic Risk Score for CAD. **UKB**: UK Biobank.

**Supplementary Figure 16: RMSE using MSGene versus FRS30**
Here, we show the RMSE overall (SEM) compared to the FRS30 year score without calibration **(A)**, FRS30RC, with calibration according to Rospleszcz et al[16] **(B)** Given that recalibration is not guaranteed to preserve the overall incidence in the population, we also performed a sensitivity analysis in which we further standardized to reflect average predictions in line with overall incidence[34] and using an additional division to match the overall incidence rate. This is for individuals progressing from the healthy state, with additional RMSE computed in supplementary table 1. In this paper, we discuss results using the traditional recalibration. In general, while further standardization improves overall RMSE, it increases the RMSE for younger individuals.
**RMSE**: Root mean squared error. **FRS30**: Framingham 30 year score. **FRS30 RC:** Framingham 30 year with recalibration according to[16], **FRS30 RC/div**: Framingham after further division to normalize overall incidence rate, in our data this was by 1.83 to normalize incidence rate to 11.1% overall.

**Supplementary Figure 17: Smoothed Fit across ages.**
We consider the unsmoothed coefficients extracted for a sample model from Health to CAD over 40 years of follow-up. We show the smoothed coefficients (‘*custom loess’*, here green) using our weighted least square regression that weights each state-state-age specific coefficient according to those within a 20-year range according to their distance and inverse variance. Here, we use polynomial degree 2, consider neighbors within 20 years and compare to a Standard loess fit (R package Stats, v 3.6.2) with span 0.75 and with (or without) weights according to inverse variance (*Standard LOESS weighted, unweighted*) for the transition from health to CAD. We provide this via a user interface: https://surbut.shinyapps.io/testapp/.
**f.31.0.01**: sex; **anti-htn now**: time-dependent anti-hypertensive use, *CAD-PRS*: Polygenic risk score.

| | Not Member (N=259287) | Member (N=221351) | Overall (N=480638) |
|---|---|---|---|
| **Sex** | | | |
| Female | 139975 (54.0%) | 120678 (54.5%) | 260653 (54.2%) |
| Male | 119312 (46.0%) | 100673 (45.5%) | 219985 (45.8%) |
| **Birthdate** | | | |
| Mean (SD) | 1950 (8.14) | 1950 (8.08) | 1950 (8.11) |
| Median [Min, Max] | 1950 [1930, 1970] | 1950 [1940, 1970] | 1950 [1930, 1970] |
| **Years Followed** | | | |
| Mean (SD) | 29.4 (8.06) | 29.5 (8.00) | 29.4 (8.03) |
| Median [Min, Max] | 30.5 [0.375, 47.6] | 30.6 [1.44, 44.5] | 30.5 [0.375, 47.6] |
| **Develop Hypertension** | | | |
| 0 | 158197 (61.0%) | 131989 (59.6%) | 290186 (60.4%) |
| 1 | 101090 (39.0%) | 89362 (40.4%) | 190452 (39.6%) |
| **Develop Coronary Disease** | | | |
| No | 231094 (89.1%) | 196084 (88.6%) | 427178 (88.9%) |
| Yes | 28193 (10.9%) | 25267 (11.4%) | 53460 (11.1%) |
| **Develop Diabetes** | | | |
| No | 234542 (90.5%) | 198400 (89.6%) | 432942 (90.1%) |
| Yes | 24745 (9.5%) | 22951 (10.4%) | 47696 (9.9%) |
| **Develop Hyperlipidemia** | | | |
| No | 199488 (76.9%) | 167556 (75.7%) | 367044 (76.4%) |
| Yes | 59799 (23.1%) | 53795 (24.3%) | 113594 (23.6%) |
| **Current Smoker** | | | |
| No | 231921 (89.4%) | 198045 (89.5%) | 429966 (89.5%) |
| Yes | 27366 (10.6%) | 23306 (10.5%) | 50672 (10.5%) |
| **Proportion White** | | | |
| Yes | 221475 (85.4%) | 95626 (88.4%) | 417101 (86.8%) |
| **Age Hypertension** | | | |
| Mean (SD) | 62.6 (11.2) | 61.9 (11.5) | 62.3 (11.3) |
| Median [Min, Max] | 63.0 [0.433, 87.0] | 62.5 [0.446, 84.3] | 62.9 [0.433, 87.0] |
| **Age CAD** | | | |
| Mean (SD) | 67.7 (8.34) | 67.5 (8.40) | 67.6 (8.37) |
| Median [Min, Max] | 68.5 [40.0, 87.0] | 68.3 [40.0, 84.3] | 68.5 [40.0, 87.0] |
| **Age Diabetes** | | | |
| Mean (SD) | 67.4 (9.25) | 67.2 (9.29) | 67.3 (9.27) |
| Median [Min, Max] | 68.6 [0.476, 87.0] | 68.4 [0.465, 84.3] | 68.5 [0.465, 87.0] |
| **Age Hyperlipidemia** | | | |
| Mean (SD) | 65.9 (8.97) | 65.5 (9.08) | 65.7 (9.02) |
| Median [Min, Max] | 66.2 [0.0137, 87.0] | 65.7 [0.0465, 84.3] | 66.0 [0.0137, 87.0] |

**Supplementary Figure 1.**

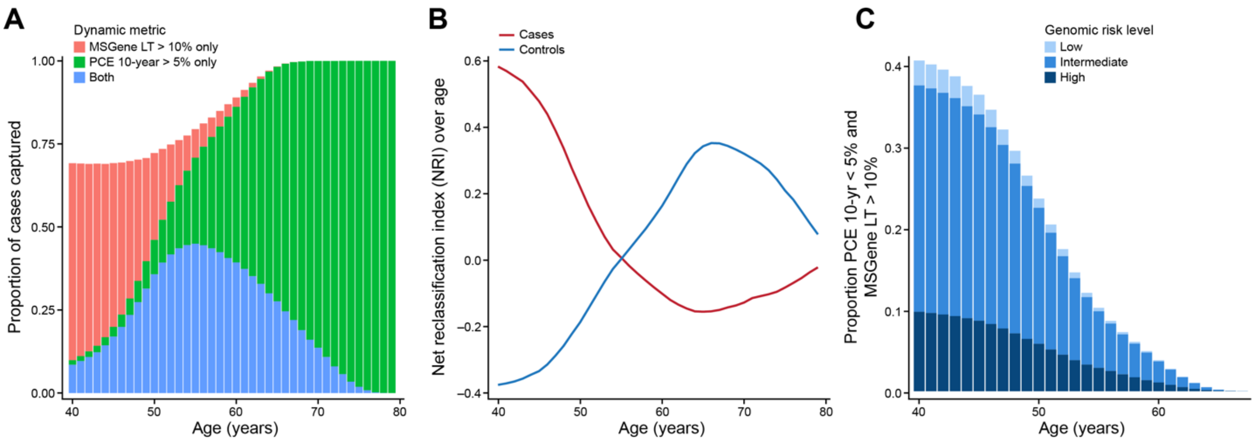

**Supplementary Figure 2.**

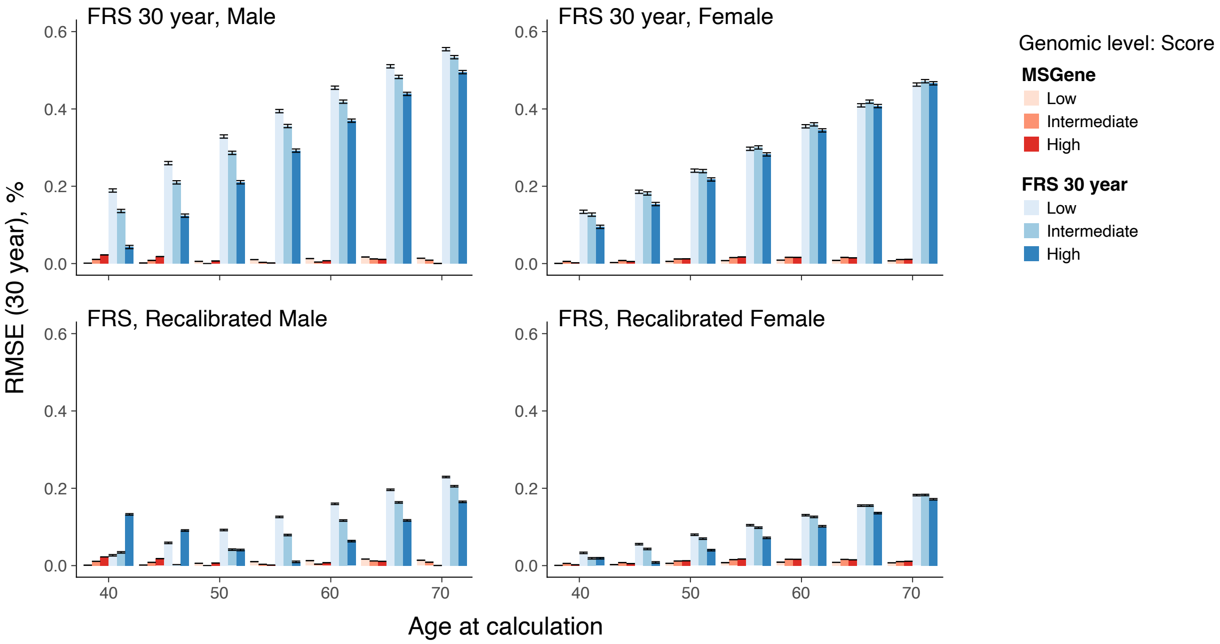

**Supplementary Figure 3.**

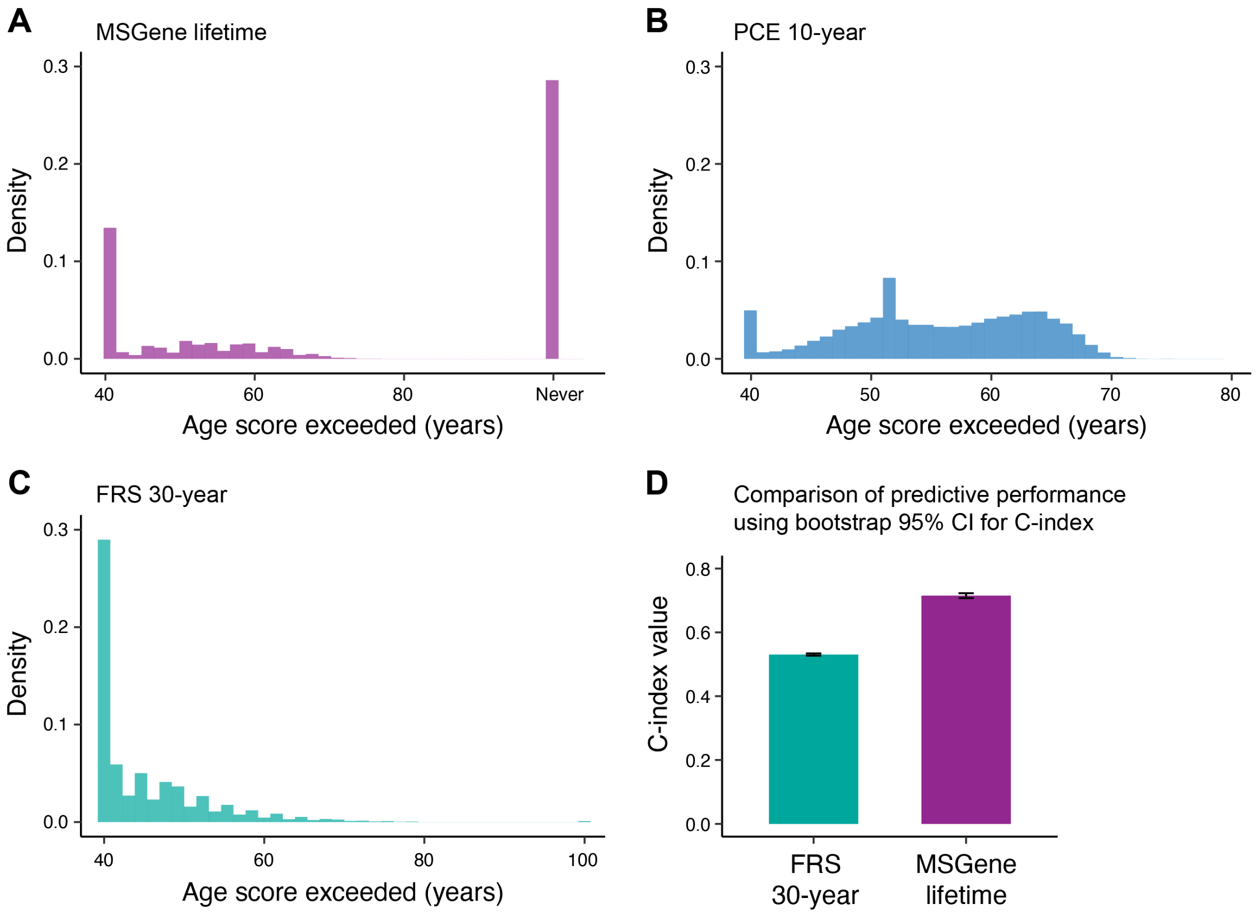

**Supplementary Figure 4.**

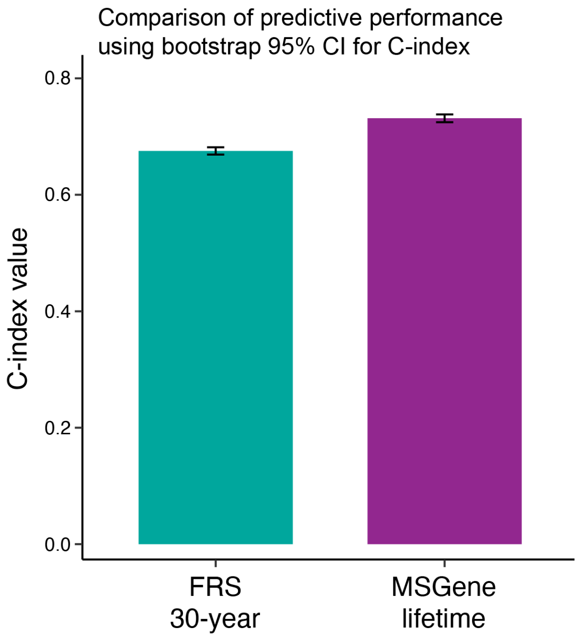

**Supplementary Figure 5.**

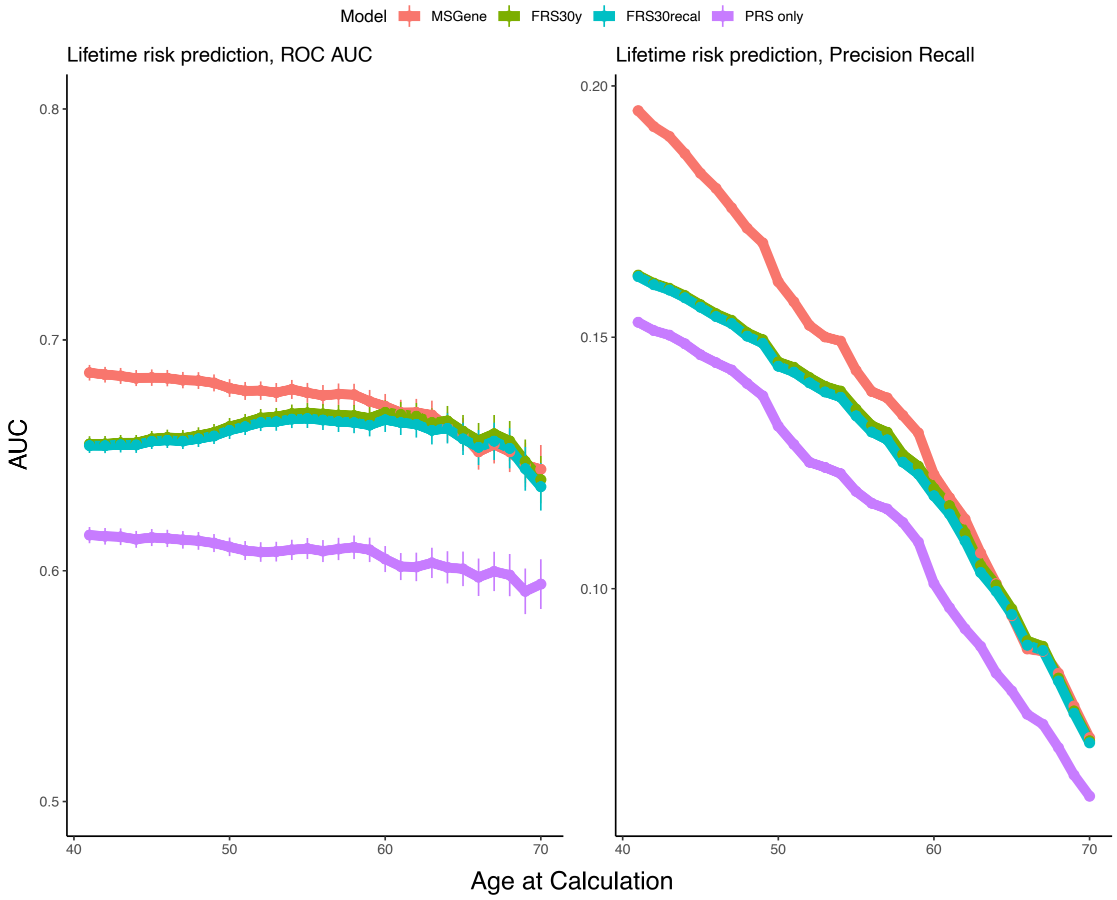

**Supplementary Figure 6.**

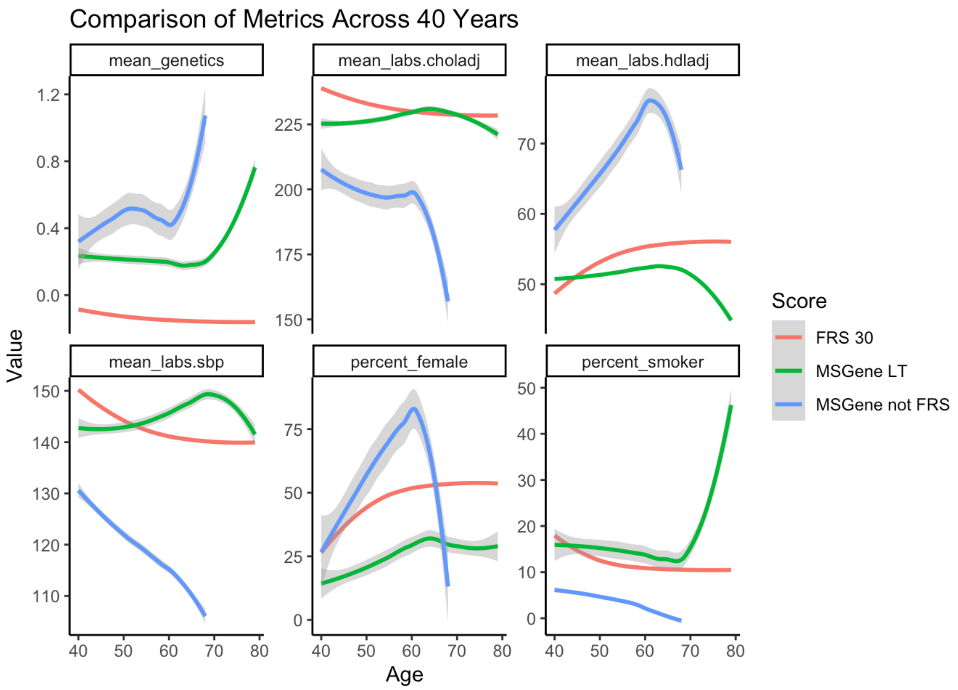

A.

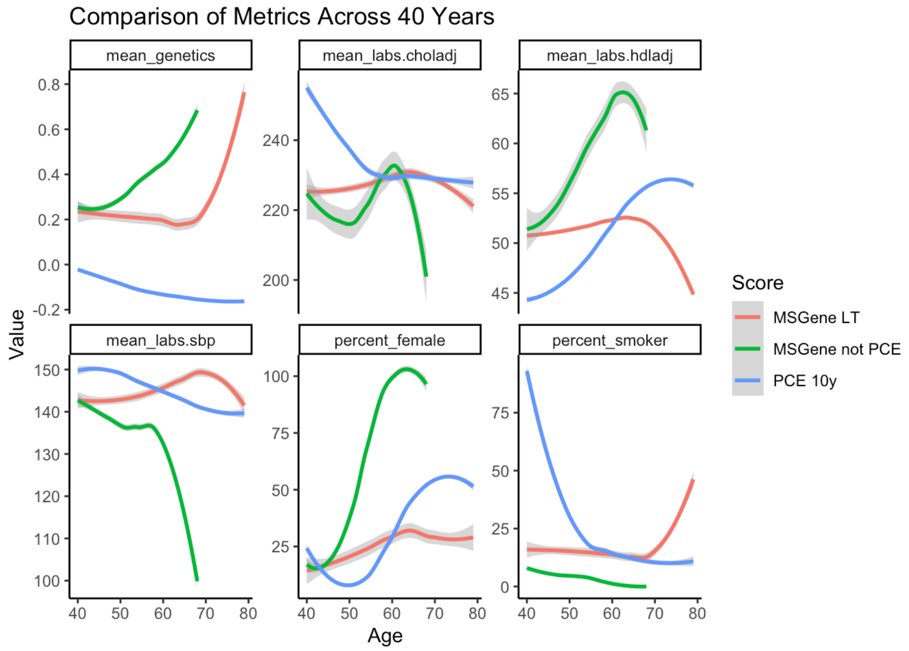

B.

**Supplementary Figure 7.**

.

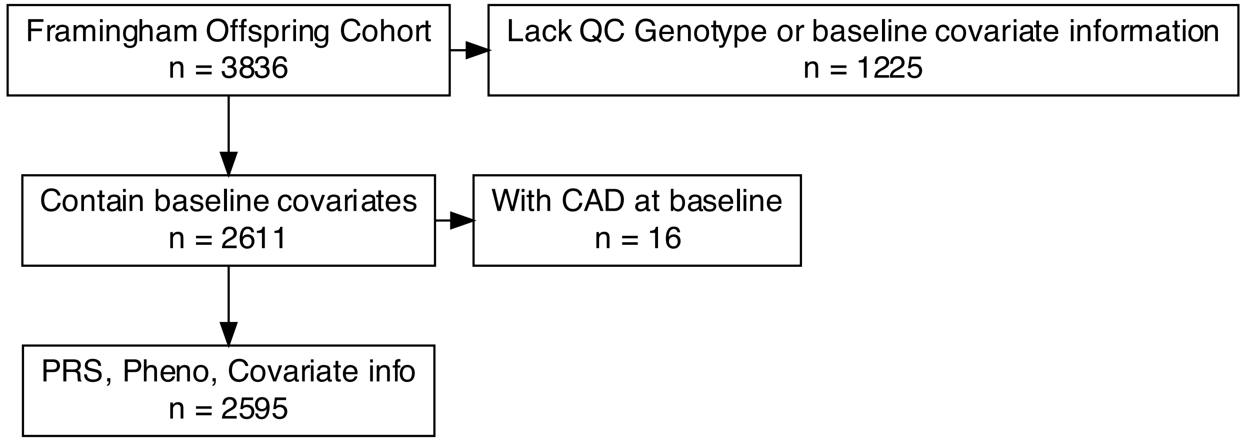

B.

| | **Low Genomic Risk (N=506)** | **Intermediate Genomic Risk (N=1575)** | **High Genomic Risk (N=514)** | **Overall (N=2595)** |
|---|---|---|---|---|
| **Sex** | | | | |
| Female Number (%) | 266 (52.6%) | 822 (52.2%) | 282 (54.9%) | 1370 (52.8%) |
| Male Number (%) | 240 (47.4%) | 753 (47.8%) | 232 (45.1%) | 1225 (47.2%) |
| **Age of First Measured** | | | | |
| Median [IQR] | 34.0 [27.0, 42.0] | 33.0 [27.0, 41.0] | 34.0 [28.0, 41.0] | 33.0 [27.0, 41.0] |
| **Develop Hypertension** | | | | |
| Mean (SD) | 0.279 (0.449) | 0.331 (0.471) | 0.358 (0.480) | 0.326 (0.469) |
| **Develop Coronary Disease** | | | | |
| Number (Percent) | 66 (13.0%) | 261 (16.6%) | 151 (29.4%) | 478 (18.4%) |
| **Develop Hyperlipidemia** | | | | |
| Mean (SD) | 0.818 (0.386) | 0.841 (0.366) | 0.891 (0.312) | 0.847 (0.360) |
| **Start an anti-Hypertensive** | | | | |
| Mean (SD) | 0.532 (0.499) | 0.630 (0.483) | 0.689 (0.463) | 0.623 (0.485) |
| **Current Smoker** | | | | |
| Mean (SD) | 0.362 (0.481) | 0.413 (0.493) | 0.416 (0.493) | 0.404 (0.491) |
| **Years Followed** | | | | |
| Mean (SD) | 36.8 (4.80) | 36.6 (5.12) | 36.5 (5.18) | 36.6 (5.07) |
| Median [Min, Max] | 38.4 [13.3, 42.1] | 38.2 [11.8, 42.3] | 38.3 [12.7, 42.3] | 38.3 [11.8, 42.3] |

**Supplementary Figure 8.**

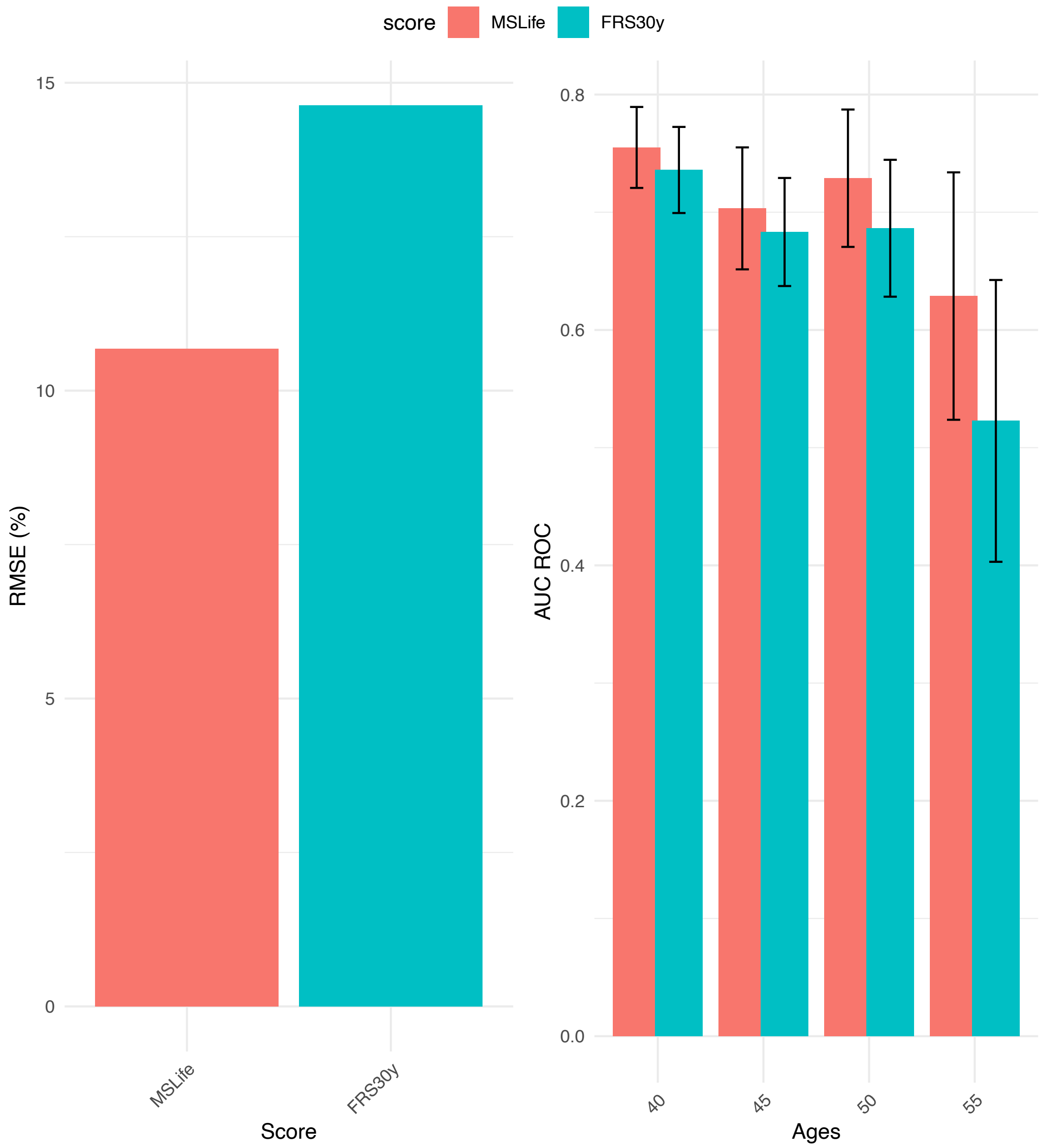

**Supplementary Figure 9.**

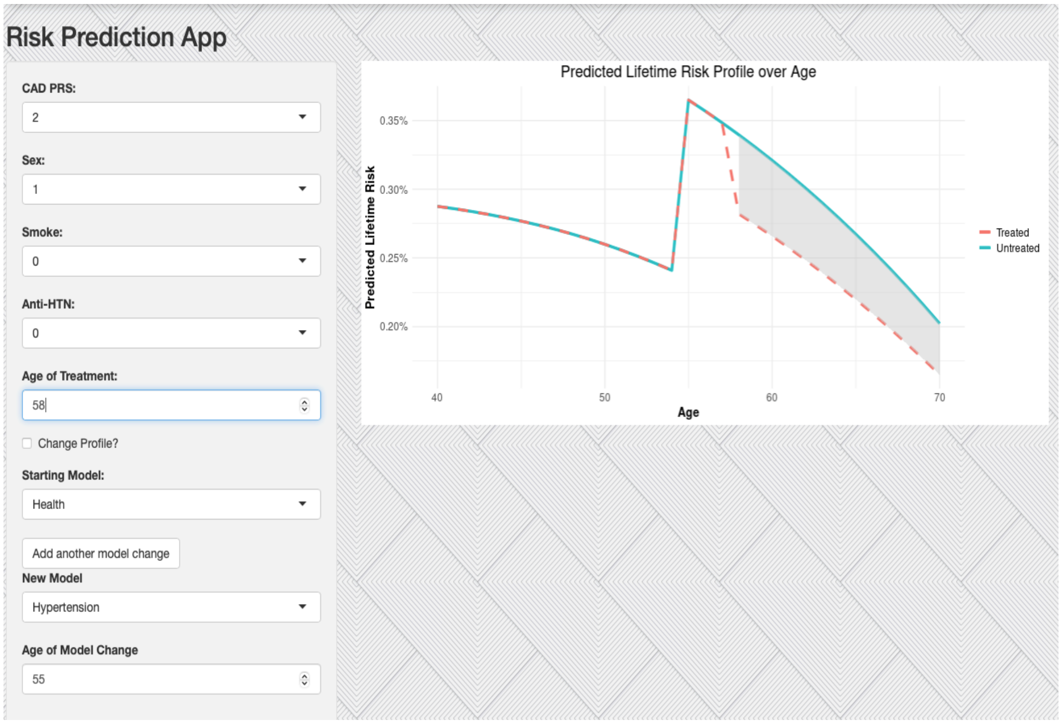

**Supplementary Figure 10.**

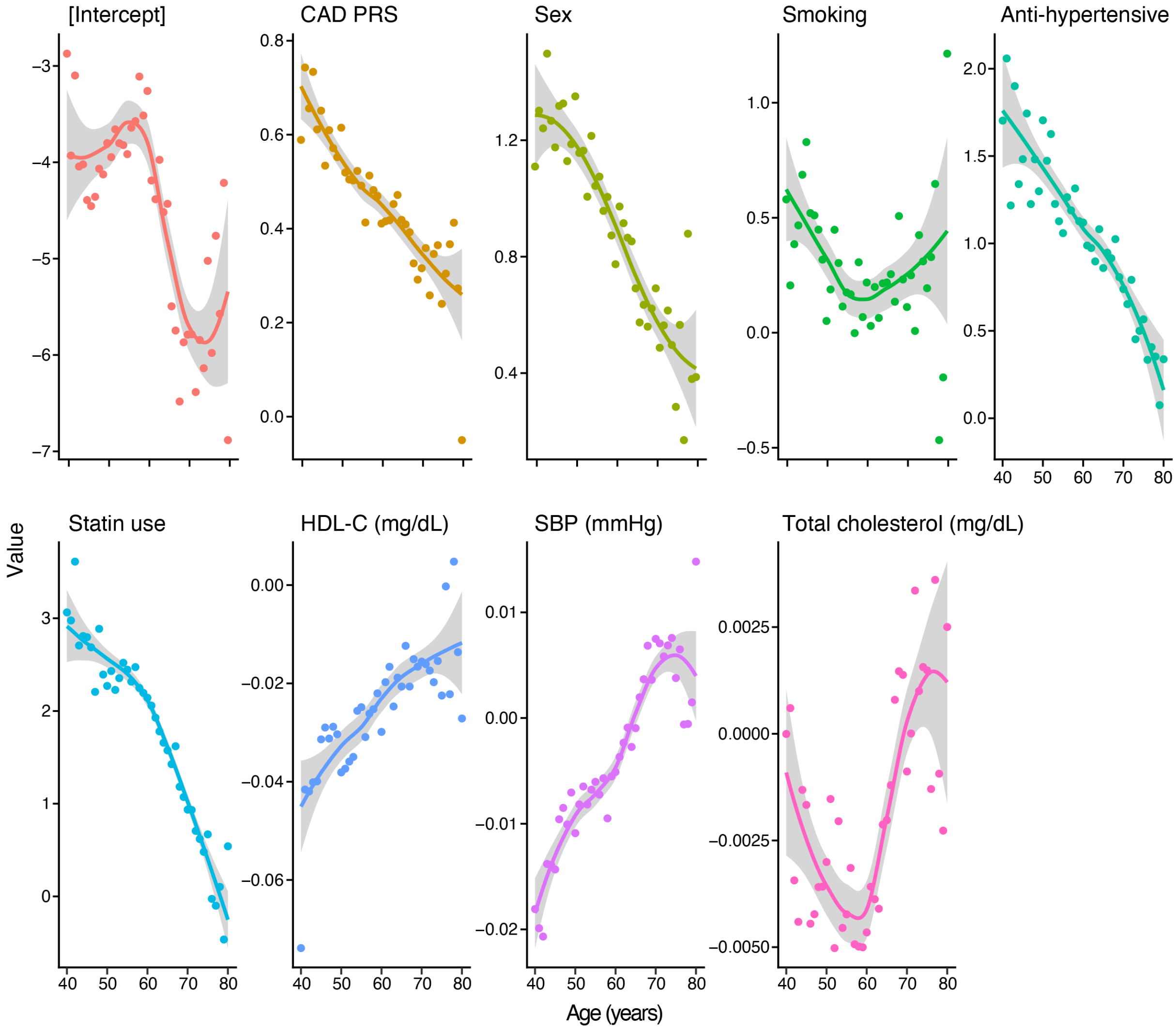

**Supplementary Figure 11.**

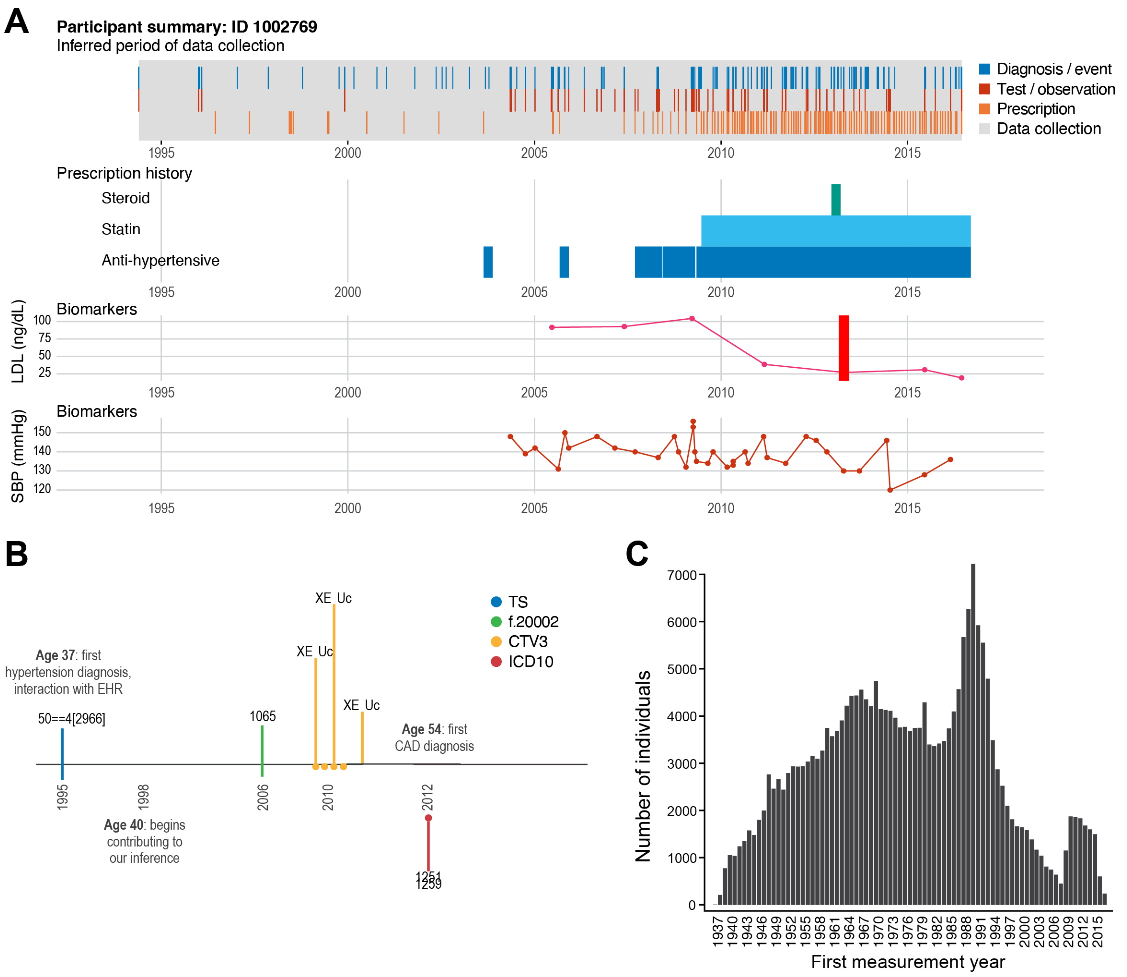

**Supplementary Figure 12.**

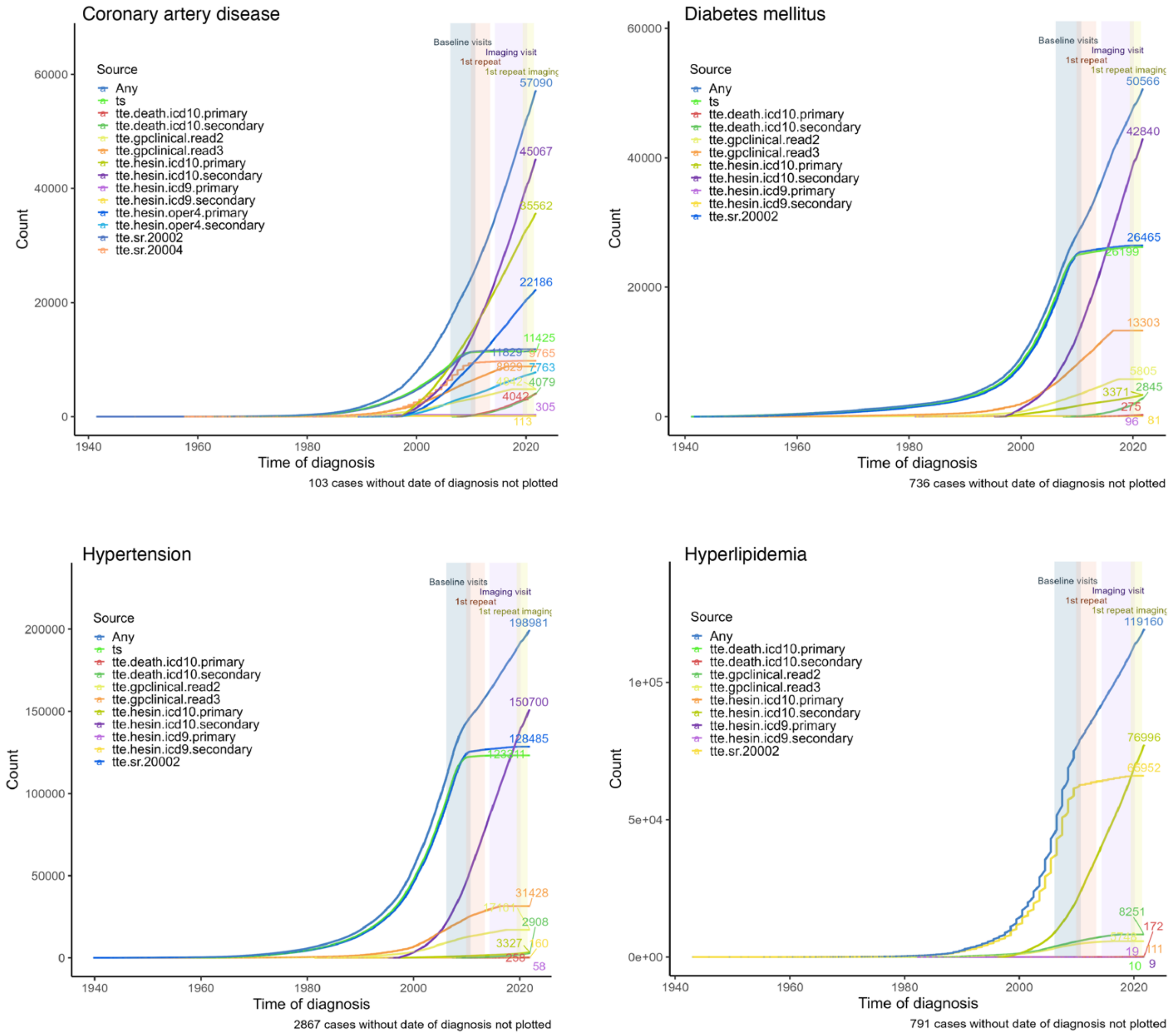

**Supplementary Figure 13.**

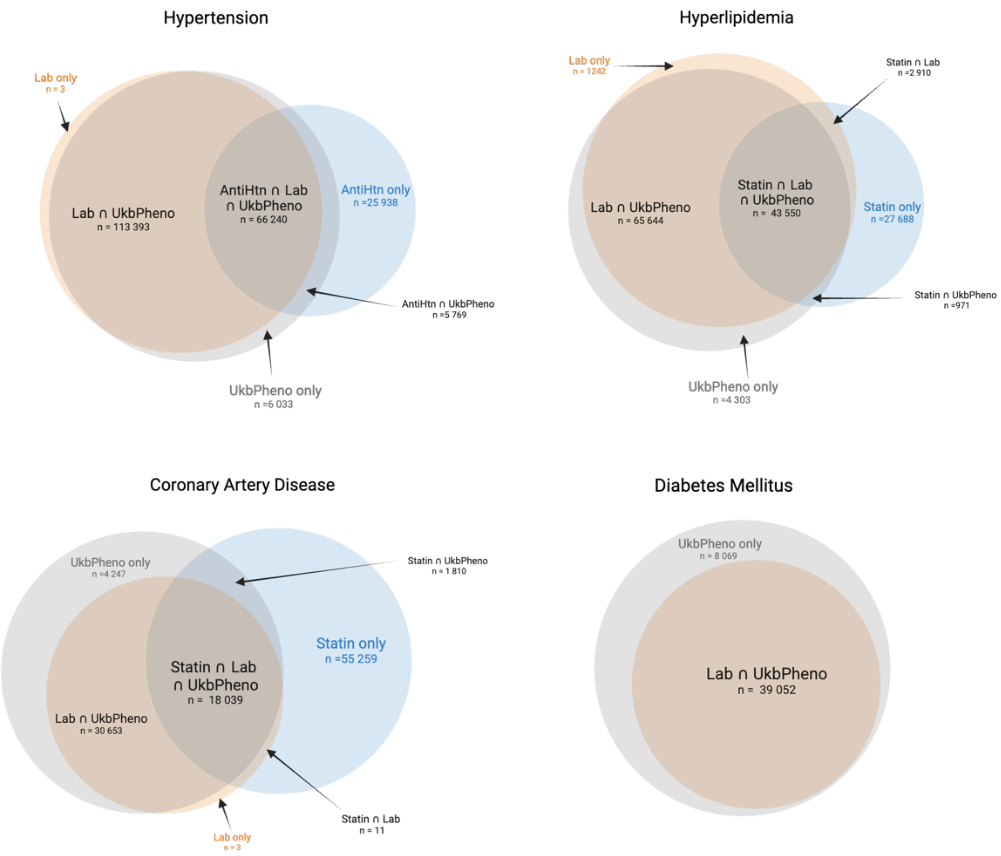

**Supplementary Figure 14.**

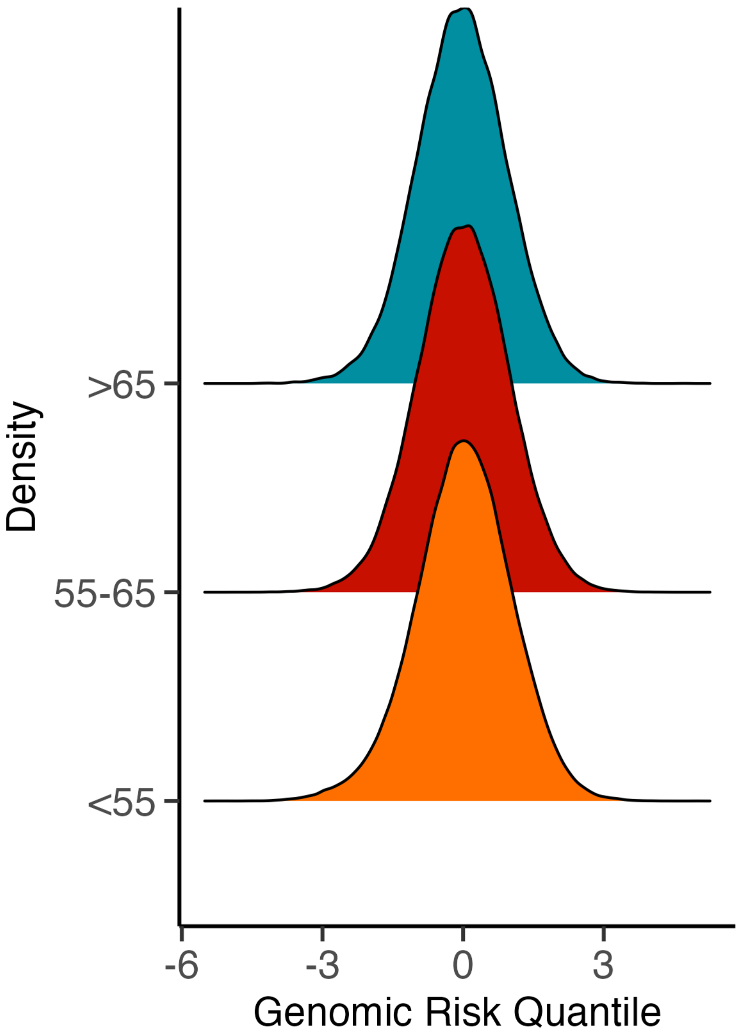

**Supplementary Figure 15.**

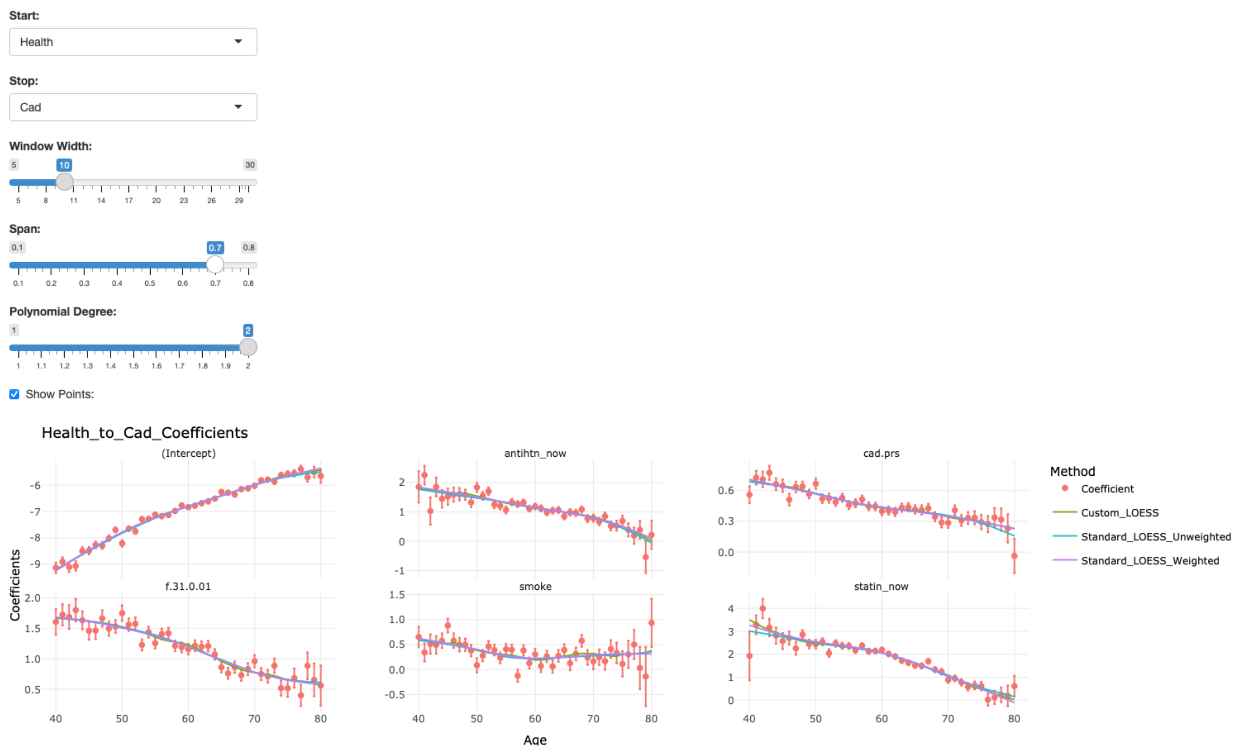

**Supplementary Figure 16.**

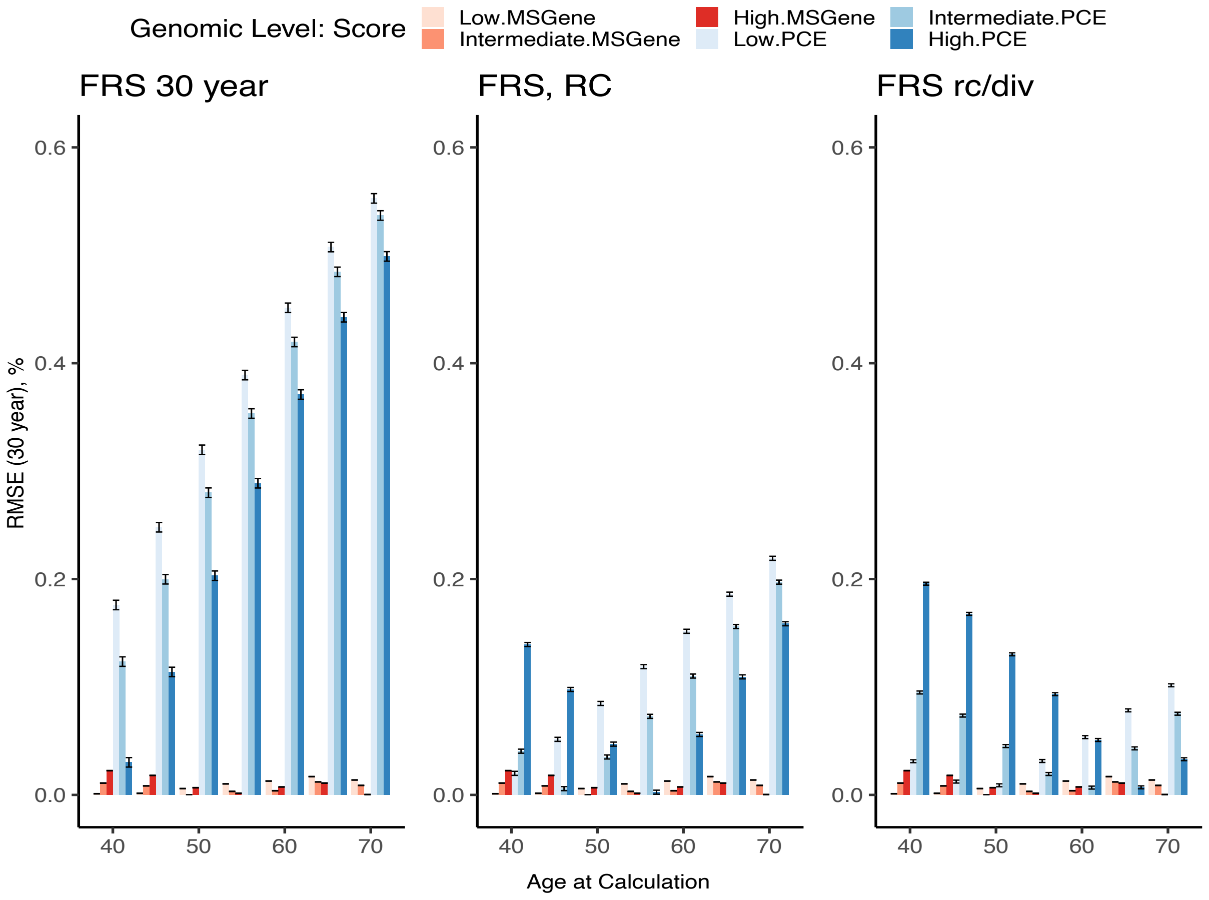

300
301
302 **Supplementary Figure 17.**
303

Additional Supplementary Materials

1. Excel Tables 2-17: Risks2-17Urbutetal.xls
2. MSGene **App**: https://surbut.shinyapps.io/risk/
3. MSGene smoothing interface: https://surbut.shinyapps.io/testapp/
4. GitHub Code for MSGene model:
   https://github.com/surbut/MSGene
5. GitHub Vignettes:
   https://surbut.github.io/MSGene/usingMarginal.html
   https://surbut.github.io/MSGene/vignette.html
